# Does home-based screening and health information provision improve hypertension diagnosis, treatment, and control? A regression discontinuity analysis in urban India

**DOI:** 10.1101/2024.02.26.24303288

**Authors:** Michaela Theilmann, Sneha Mani, Pascal Geldsetzer, Shivani A. Patel, Mohammed K. Ali, Harsha Thirumurthy, KM Venkat Narayan, Viswanathan Mohan, Dorairaj Prabhakaran, Nikhil Tandon, Nikkil Sudharsanan

**Author notes:** Corresponding author: Michaela Theilmann, Im Neuenheimer Feld 130.3, 69120 Heidelberg, Germany. Joint first-authors, equal contribution (Michaela Theilmann, Sneha Mani).

## Abstract

**Background:** In India, several state governments are implementing or considering home-based hypertension screening programs to improve population-wide diagnosis and blood pressure (BP) control rates. However, there is limited evidence on the effectiveness of home-based screening programs in India.

**Methods:** Using six waves of population-representative cohort data (N = 15,573), we estimate the causal effect of a home-based hypertension screening intervention on diagnosis, treatment, and BP using a novel application of the Regression Discontinuity Design.

**Findings:** We find that screening individuals’ BP in their homes and providing health information and a referral to those with elevated BP did not meaningfully improve hypertension diagnosis (0.1, p-value: 0.82), treatment (−0.2, p-value: 0.49), or BP levels (systolic: −1.8, p-value: 0.03; diastolic: 0.5, p-value: 0.39). This null effect is robust across subpopulations and alternative specifications.

**Interpretation:** Our findings suggest that a lack of knowledge of one’s hypertension status might not be the primary reason for low diagnosis and treatment rates in India, where other structural and behavioral barriers may be more relevant. Adapting screening efforts to address these additional barriers will be essential for translating India’s screening efforts into improved population health.

**Funding:** This study received no funding.

**Research in context: Evidence before this study:**

- Indian state governments are implementing and scaling-up large home-based screening programs to address the growing burden of cardiometabolic diseases
- Studies evaluating home-based screening activities in China and Malawi find that they lead to modest improvements in blood pressure levels
- However, studies from South Africa and Germany find null effects of home-based screening on blood pressure and long-term cardiometabolic outcomes

**Added value of this study:**

- We provide the first evaluation of home-based hypertension screening in India using data representative of adults aged 30 years and older in two of India’s largest cities.
- In addition to blood pressure level, we investigate the effect of screening on diagnosis and treatment initiation to identify the effects of screening across the continuum of care.
- We find that home-based hypertension screening did not improve hypertension outcomes suggesting that a lack of knowledge of one’s hypertension status is not the main barrier to health care seeking and achievement of hypertension control.

**Implications of all the available evidence:**

- The mere provision of information on an individual’s hypertension status does not seem to increase hypertension diagnosis and treatment initiation.
- Other barriers, such as health literacy and inadequate communication strategies, need to be considered when designing home-based hypertension screening programs.
- Before scaling up existing home-based screening programs, health communication needs to be adapted to local needs and their effectiveness evaluated.

## 1 Introduction

Uncontrolled hypertension substantially increases the risk of cardiovascular diseases such as heart attack and stroke and is the leading risk factor for mortality worldwide. ^1^ Controlling hypertension is a highly cost-effective way of reducing premature mortality and is considered a global best buy. ^2–4^ Unfortunately, in many countries, including India, the prevalence of uncontrolled hypertension is high while rates of hypertension diagnosis and treatment are low. In 2019-2021, an estimated 28% of adults aged 18 or older in India had hypertension, yet just 37% of these individuals were diagnosed. Additionally, only 45% among the diagnosed reported taking medication, and of these treated individuals, only 53% had their blood pressure (BP) under control. ^5^ Due to population aging, the number of people in need of hypertension care in India is expected to more than double over the coming decades making the improvement of hypertension control an urgent health priority. ^6^

To address the growing burden of cardiometabolic diseases, the Indian government initiated “The National Programme for Prevention and Control of Cancer, Diabetes, Cardiovascular Disease and Stroke” in 2010, followed by the “National Multisectoral Action Plan for Prevention and Control of Common Non-Communicable Diseases 2017-2022” in 2013, and the “National Programme for Prevention and Control of Non-Communicable Diseases” in 2023. ^7–9^ A key component of all three plans is population- and community-based hypertension screening and several state governments have started to implement home-based screening initiatives. ^8,10^ The main theory of change behind such home-based screening efforts, which can be considered “information intervention”, ^11–13^ is that making individuals aware that they may have hypertension and referring them to the health system for follow-up will encourage individuals to seek formal health care, receive a diagnosis, initiate treatment, and ultimately control their BP. These interventions thus assume that a lack of awareness about one’s hypertension status and care needs is the critical barrier to achieving BP control. Yet the limited available evidence from China, Malawi, and South Africa evaluating home-based screening initiatives show that it has small or even null effects on BP levels, ^14–16^ suggesting that many individuals may not seek a confirmatory diagnosis or initiate treatment after being told they should seek further care.

While informative, the existing evidence has two primary limitations for informing policy in the Indian context. First, none of the above studies used data from India, which limits their generalizability to the Indian context. Second, although all three studies estimated the effect of home-based screening on BP levels, only one evaluated the effect of screening on hypertension diagnosis and treatment initiation, which are prerequisites for achieving BP control. Investigating these intermediate care continuum steps is essential for identifying at what stage linkage to care fails and where additional interventions are needed.

We address these important limitations and evaluate a home-based hypertension screening, information provision, and referral on confirmatory diagnosis, drug treatment, and BP in urban India. Our study leverages a quasi-experimental design with rich longitudinal cohort data with measured BP information. We also investigate whether the screening and information intervention has differential effects based on individuals’ education, age, prior diagnoses of other cardiometabolic diseases, and city of residence. Overall, our results reveal the extent to which home-based screening has been effective in urban India and help inform ongoing efforts to implement and expand home- and community-based screening in India and similar contexts.

## 2 Data and Methods

### 2.1 Data

We used data from all six waves of the Centre for Cardiometabolic Risk Reduction in South Asia (CARRS) cohort in India ^17^. CARRS is a longitudinal cohort survey conducted between 2010 and 2018 (wave 1: October 2010 to November 2012; wave 2: November 2011 to March 2013; wave 3: March 2013 to April 2014; wave 4: February 2014 to June 2014; wave 5: January 2016 to February 2017; wave 6: January 2017 to April 2018) in Chennai and Delhi, two of the largest cities in India with populations of 4.6 and 16.8 million, respectively ^18^.

The survey employed a multi-stage cluster random sampling design yielding a sample representative of the population aged 20 years or older in both cities. First, twenty municipal wards from Delhi and Chennai each were randomly selected. Next, five census enumeration blocks were randomly selected from each ward, and a household listing was conducted in each block. In each block, 20 households were randomly selected and one male and one female household member aged 20 years or older were randomly selected from each household. ^17^ Individuals were approached and invited to participate in the survey in all six waves.

### 2.2 Blood Pressure Measurement

The CARRS study measured BP as part of routine data collection in wave 1 (2010-2012), wave 3 (2013-2014), and wave 5 (2016-2017). Trained field staff recorded at least two BP measurements toward the end of the interview using an Automated Omron HEM-7080 or HEM-708016 device ^19,20^. Participants had to rest for five minutes before the first measurement was taken. A second measurement was taken at least 30 seconds after the first measurement. A third measurement was taken if the difference between measurements was greater than 10 mmHg for systolic BP or 6 mmHg for diastolic BP.

### 2.3 Screening Intervention

The main intervention action we evaluate is the effect of informing and referring potentially hypertensive individuals to the formal healthcare system for diagnosis and treatment initiation. Specifically, during the household visit, if an individual’s average systolic BP was *≥* 140 mmHg or average diastolic BP was *≥* 90 mmHg, the field staff informed the individual that they might have hypertension, provided basic information on the importance of hypertension control, and instructed them to visit a healthcare facility for a formal assessment and diagnosis. The field staff’s activities are, therefore, comparable to a community-based screening intervention, in which screeners visit people’s homes, measure their BP, and refer those with high BP for formal care.

### 2.4 Outcomes

We investigated four outcomes representing the different stages of the hypertension care cascade. The first outcome is whether an individual obtained a confirmatory diagnosis of hypertension after community hypertension screening. This is measured by an individual’s self-reported physician diagnosis of hypertension in the past 12 months. The second outcome is whether an individual initiated hypertension medication following the home-based screening visit. This is measured by an individual’s self-report of currently taking allopathic medication for high BP. This indicator is conditional on an individual’s self-report of a confirmatory hypertension diagnosis in the past 12 months and should, thus, be interpreted as “received confirmatory diagnosis in past 12 months and currently takes hypertension medication”. Table S1 presents the exact wording used in the questionnaire to measure these two outcomes. The third and fourth outcomes are average systolic and diastolic BP at follow-up.

### 2.5 Analysis Samples

BP was only measured in waves 1, 3, and 5 of the 6 total CARRS survey waves. Thus, for the confirmatory diagnosis and drug treatment outcomes, we created three baseline-follow-up pairs of data (waves 1 to 2, waves 3 to 4, and waves 5 to 6). In each pair, the BP measurement came from the baseline wave, while the outcome data on diagnosis and treatment was from the follow-up wave. We pooled data from the three pairs to construct the analytical dataset for these two outcomes. For the systolic and diastolic BP outcomes, we could only use waves with BP measurements and, thus, created two baseline-follow-up pairs (waves 1 and 3, and waves 3 and 5), which we pooled for the analysis.

We had three separate analysis samples: one for the diagnosis outcome, one for the treatment outcome, and one for the systolic and diastolic BP outcomes. For the diagnosis sample, we restricted our data to individuals who were aged 30+ in the baseline year - which mirrors the age range targeted by the Indian government’s NP-NCD ^9^ - and who did not report a diagnosis of hypertension at the baseline or prior waves. For the treatment sample, we restricted our data by age and to those who did not report currently taking treatment in the baseline wave. For the BP outcomes, we only restricted the sample to individuals aged 30 years or older.

### 2.6 Methods and Identification Strategy

Since the referral and information provision activities were not randomized, we could not evaluate them using standard randomized trial methods and rather needed to employ an alternative approach to estimate causal effects. We employed the sharp regression discontinuity design (RDD) to evaluate the impact of home-based hypertension screening on confirmatory hypertension diagnosis, drug treatment initiation, and BP levels. The RDD is a quasi-experimental method that allows for the identification of a causal effect in situations where randomization was not possible or not done but individuals were allocated to intervention or control groups based on a clear, pre-specified rule. ^21–25^ This rule relies on the measurement of a continuous variable known as the running variable. In the context of our analysis, the intervention consisted of informing participants with high BP that they may have hypertension and referring them to a healthcare facility. The intervention was assigned based on the continuous running variable of BP with the rule that only those with systolic BP *≥* 140 mmHg or diastolic BP *≥* 90 mmHg should receive the intervention. Survey participants with a systolic BP below 140mmHg and a diastolic BP below 90mmHg did not receive such a referral and thus no intervention.

The traditional RDD relies on one running variable and one threshold. However, in this study, individuals were referred if either their systolic or diastolic BP was high. For this reason, we standardized and combined the two measurements into one running variable, resulting in one threshold. ^26,27^ First, we standardized each value by subtracting the threshold value (140 for systolic and 90 for diastolic BP) from an individual’s average measurement result and then dividing it by the sample’s standard deviation. This resulted in an individual’s standardized distance from the threshold for the systolic and diastolic BP measurements, respectively. In the second step, we chose the larger distance to define where an individual was located relative to the threshold. In summary, we applied the following equation:

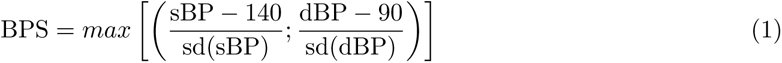

with BPS = standardized BP as the running variable, sBP = systolic BP, dBP = diastolic BP, and sd = standard deviation.

If BPS *<* 0, the individual was below the threshold and did not receive the intervention. If BPS *≥* 0, the individual was at or above the threshold and received the intervention. As the calculation of the running variable depends on the distribution of the underlying data, we generated a separate running variable for the four outcomes overall and separated by sociodemographic characteristics.

The main identifying assumption required for the RDD to produce causal estimates is that in the absence of the intervention, the relationship between the running variable (standardized BP) and the outcomes (hypertension diagnosis, drug treatment, and BP levels) does not change discontinuously at the threshold. Thus, any jump in the outcome around the threshold value represents the causal effect of the intervention. Heuristically, this assumption is often described as “individuals just above and below the threshold are identical except that those above the threshold received the intervention”. In the context of our study, this assumption is highly plausible as there is no physiological change that occurs at the 140/90 threshold that would induce individuals to seek care beyond the referral from the survey team.

In our analysis, we followed the standard RDD methodology described in Cattaneo et al. (2019). ^26^ Specifically, we estimated a model with local linear trends, triangular kernel weights, and a mean square error (MSE) optimal bandwidth. This bandwidth determines which range of BP values the RDD is estimated on and was used for the point estimates and inference. The calculation of the MSE optimal bandwidth is driven by the data and, thus, prevents arbitrary definition or manipulation by researchers. Furthermore, we included a time trend by adding survey pair dummies and clustered standard errors at the individual level. The point estimate is interpreted as the treatment effect on individuals who are directly at the cut-off (i.e. with BPS = 0), which is the local average treatment effect (LATE).

Before conducting the formal RDD analysis, we first plotted the standardized BP measurement against the respective outcome for observations within the MSE optimal bandwidth. Then, we fitted the local linear trend to assess whether the intervention might have had an effect.

Analyses were conducted in R version 4.0.3 using the rdrobust package.

### 2.7 Heterogeneity analyses

We also investigated whether the screening information had differential effects across population sub-groups. To do so, we estimated separate RDD models by sex (male, female), education categories (no education or incomplete primary school (0-4 years), primary school completed (5-9 years), and high school or more completed), age group (30-39,40+), by self-report of prior cardiometabolic disease diagnoses (diabetes, stroke, or heart attack), and by city of residence (Chennai, Delhi).

## 3 Results

### 3.1 Sample Description

Pooling observations from all three survey pairs yielded data on 27,554 observations of adults (100%) aged 30 years or older at baseline (Figure 1). We excluded 6,238 individuals (23%) who did not participate in the follow-up survey and 990 (4%) because they did not have a valid BP reading. For the confirmatory hypertension diagnosis outcome, 615 (2%) observations were excluded because they did not have information on hypertension diagnosis at the baseline or follow-up. This resulted in 19,711 (72%) observations with the information required to analyze the diagnosis outcome. Subsequently, we excluded the 4,138 individuals who reported a previous hypertension diagnosis at the baseline or any earlier wave, resulting in a sample of 15,573 individuals.

**Figure 1:**
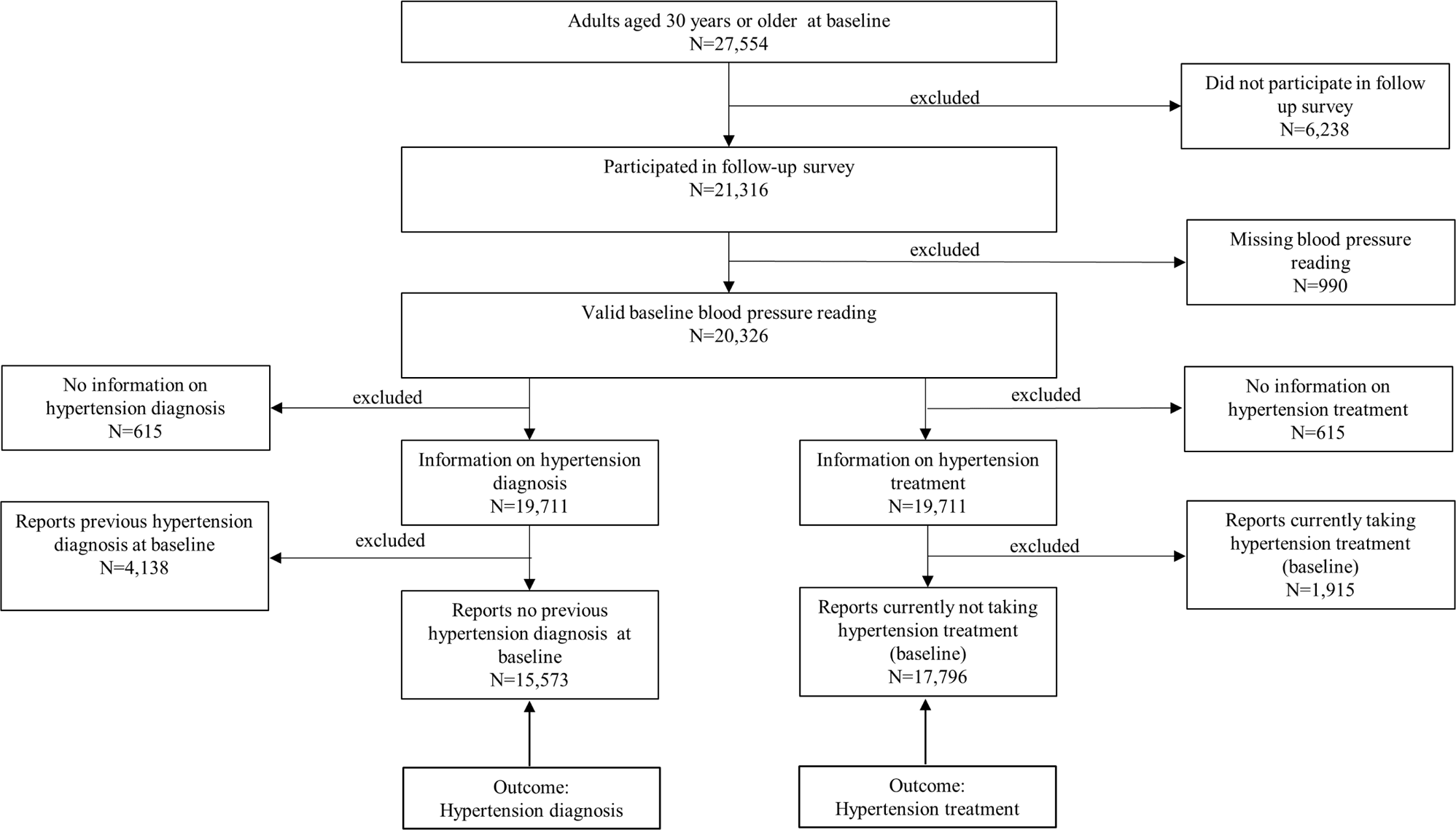
Number of observations included in analysis

For the hypertension treatment outcome, 615 (2%) did not have information on hypertension treatment at the baseline or follow up, leaving 19,711 individuals (72%) with all the information required for the analysis of the treatment outcome. Subsequently, we excluded the 1,195 individuals who reported being on medication at the baseline, resulting in a sample of 17,796 adults.

For systolic and diastolic BP, we pooled data from the two survey pairs including 18,721 adults aged 30 years and older. Of those, 13,590 (73%) had BP measurements at the respective baseline and follow-up surveys.

Tables 1, 2, and S2 display the sample characteristics for the total population as well as the male and female subpopulations. The “overall” sample consists of all observations that were included in the study. The “analytical” sample includes only observations within the MSE optimal bandwidth of the respective RDD model estimation. For all outcomes, characteristics were comparable across samples, for the total population, females, and males, and when comparing the overall and analytical samples. One notable difference was that the share of women aged 40 years or older was higher in the analytical than in the overall samples.

**Table 1:** Characteristics of the overall and analytic samples for adults ages 30 and older, hypertension diagnosis samples.

| Characteristic | Total |  | Female |  | Male |  |
| --- | --- | --- | --- | --- | --- | --- |
|  | Overall | Analytical | Overall | Analytical | Overall | Analytical |
| Number | 15,573 | 7,977 | 8,122 | 3,347 | 7,451 | 4,312 |
| Mean age, years (SD) | 46 (11) | 46 (11) | 44 (10) | 46 (10) | 47 (12) | 47 (11) |
| Age groups |  |  |  |  |  |  |
| < 40 years | 33.9% | 29.9% | 37.1% | 29.0% | 30.4% | 29.8% |
| ≥ 40 years | 66.1% | 70.1% | 62.9% | 71.0% | 69.6% | 70.2% |
| Religion |  |  |  |  |  |  |
| Hindu | 80.3% | 79.6% | 81.0% | 80.3% | 79.6% | 79.2% |
| Muslim | 10.2% | 10.9% | 9.6% | 10.3% | 11.0% | 11.3% |
| Others | 9.4% | 9.6% | 9.4% | 9.4% | 9.4% | 9.5% |
| Education categories |  |  |  |  |  |  |
| Primary | 17.6% | 16.9% | 22.7% | 23.7% | 12.1% | 11.2% |
| Secondary | 34.2% | 33.6% | 36.2% | 37.2% | 31.9% | 31.0% |
| Tertiary | 48.2% | 49.5% | 41.0% | 39.1% | 56.0% | 57.9% |
| Reported diabetes diagnosis | 12.6% | 14.1% | 12.3% | 15.4% | 12.9% | 13.6% |
| Reported prior heart attack | 2.1% | 2.0% | 1.1% | 1.1% | 3.1% | 2.7% |
| Reported prior stroke | 0.9% | 0.8% | 0.4% | 0.5% | 1.4% | 1.2% |

Notes: The table displays the characteristics of the sample used in the analysis with hypertension diagnosis as the outcome. The overall sample consists of all individuals meeting the eligibility criteria. The analytical sample consists of those individuals with a standardized blood pressure within the mean squared error optimal bandwidth of the main analysis.
Abbreviation: SD = standard deviation.

**Table 2:** Characteristics of the overall and analytic samples for adults ages 30 and older, hypertension treatment samples.

| Characteristic | Total |  | Female |  | Male |  |
| --- | --- | --- | --- | --- | --- | --- |
|  | Overall | Analytical | Overall | Analytical | Overall | Analytical |
| Number | 17,796 | 8,456 | 9,408 | 4,266 | 8,388 | 5,072 |
| Mean age, years (SD) | 46 (11) | 47 (11) | 45 (11) | 47 (11) | 48 (12) | 47 (12) |
| Age groups |  |  |  |  |  |  |
| < 40 years | 31.3% | 27.1% | 34.0% | 26.4% | 28.4% | 28.2% |
| ≥ 40 years | 68.7% | 72.9% | 66.0% | 73.6% | 71.6% | 71.8% |
| Religion |  |  |  |  |  |  |
| Hindu | 80.2% | 79.6% | 80.5% | 79.5% | 79.9% | 79.6% |
| Muslim | 10.3% | 10.7% | 9.9% | 10.5% | 10.6% | 10.8% |
| Others | 9.5% | 9.7% | 9.6% | 10.0% | 9.5% | 9.6% |
| Education categories |  |  |  |  |  |  |
| Primary | 18.1% | 17.4% | 23.7% | 24.5% | 11.7% | 10.9% |
| Secondary | 33.5% | 33.0% | 35.2% | 35.5% | 31.5% | 30.6% |
| Tertiary | 48.4% | 49.6% | 41.0% | 40.0% | 56.7% | 58.5% |
| Reported diabetes diagnosis | 14.7% | 16.5% | 14.6% | 17.7% | 14.8% | 15.4% |
| Reported prior heart attack | 2.7% | 2.5% | 1.7% | 1.7% | 3.8% | 3.3% |
| Reported prior stroke | 1.4% | 1.3% | 0.9% | 1.0% | 1.9% | 1.6% |

Notes: The table displays the characteristics of the sample used in the analysis with hypertension treatment as the outcome. The overall sample consists of all individuals meeting the eligibility criteria. The analytical sample consists of those individuals with a standardized blood pressure within the mean squared error optimal bandwidth of the main analysis.
Abbreviation: SD = standard deviation.

### 3.2 Effect of the Home-Based Screening Intervention on Confirmatory Hypertension Diagnosis

Across the baseline levels of BP within the MSE optimal bandwidth, individuals reported low levels of diagnosis in the follow-up wave (ranging from 0.0% to 11.1% in the total population, from 0.0% to 15.4% among females, and from 0.0% to 14.3% among males [Figure 2]). At the threshold used to assign the screening intervention, we find visual evidence of a small positive effect of the intervention on confirmatory diagnosis among females, and a small negative effect among males.

**Figure 2:**
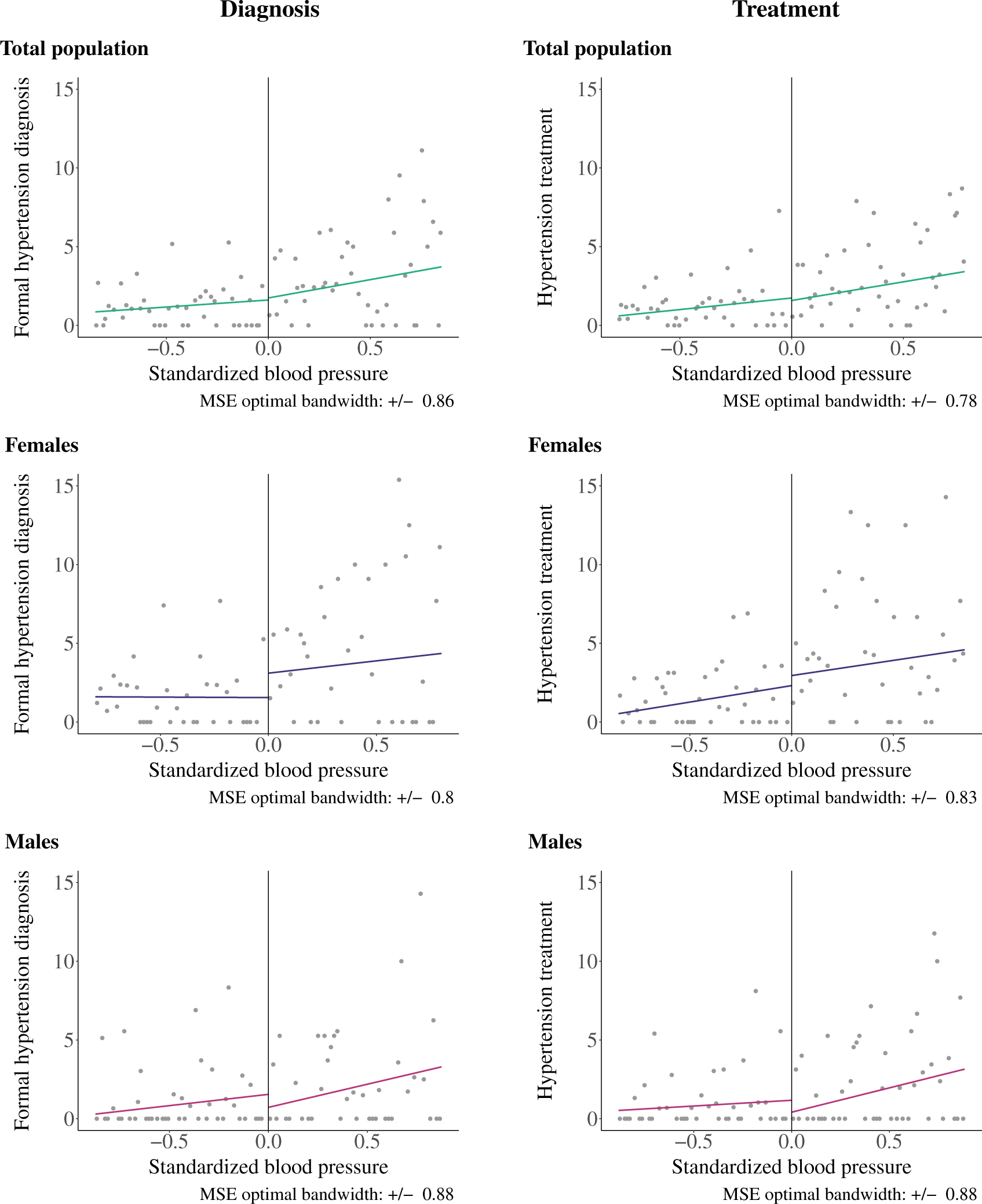
Probability of a hypertension diagnosis (left panel) and hypertension treatment initiation (right panel) among adults aged 30 and older Note: These plots only show observations within each mean squared error optimal bandwidth. The vertical line indicates the threshold set at a standardized blood pressure of zero. The colored lines represent the linear trends fitted separately for individuals with a standardized blood pressure below zero and those with a standardized blood pressure of zero or higher.

The results of the formal RDD estimation support the visual results (Table S3). We do not find evidence that the screening intervention improved diagnosis in the total population, with an estimated near-zero effect (0.1pp, 95% CI −1.4 - 1.8). While the visual results suggested a small positive effect among females and a small negative effect among males, both of these estimated effects are near zero in magnitude and estimated with confidence intervals that overlap the null (females: 1.55pp, 95% CI −1.7 - 4.4; males: −0.8pp, 95% CI −2.5 - 0.8).

### 3.3 Effect of the Home-Based Screening Intervention on Hypertension Drug Treatment

The overall levels of reported treatment were slightly lower within the MSE optimal bandwidth (ranging from 0.0% to 8.7% in the total population, from 0.0% to 28.6% among females, and from 0.0% to 11.8% among males) compared to self-reported diagnosis (Figure 2). We find a similar set of null results for the effect of the screening intervention on hypertension drug treatment, with little visual evidence and near-zero estimated effects in the total population and female and male sub-populations (Figure 2 and Table S3).

### 3.4 Effect of the Home-Based Screening Intervention on Systolic and Diastolic Blood Pressure

Similar to the results for diagnosis and treatment, we also fail to find evidence that the home-based hypertension screening intervention had an effect on either systolic or diastolic BP in any of the three populations (Figure S1, Table S4).

### 3.5 Differential Effects by Education, Age, Prior Diagnoses of Cardiovascular Disease, and City

Consistent with the overall results, we do not find evidence that the screening intervention had differential effects on confirmatory diagnosis, drug treatment, or systolic or diastolic BP across education and age groups (Figures 3 and S2, Tables S5 to S8). We find suggestive evidence of small confirmatory diagnosis and drug treatment effects among women with greater than high-school education (diagnosis: 4.7pp, 95% CI −2.0 - 7.0; treatment: 3.07pp, 95% CI −4.17 - 5.16) but are unable to statistically distinguish these effects from the null.

**Figure 3:**
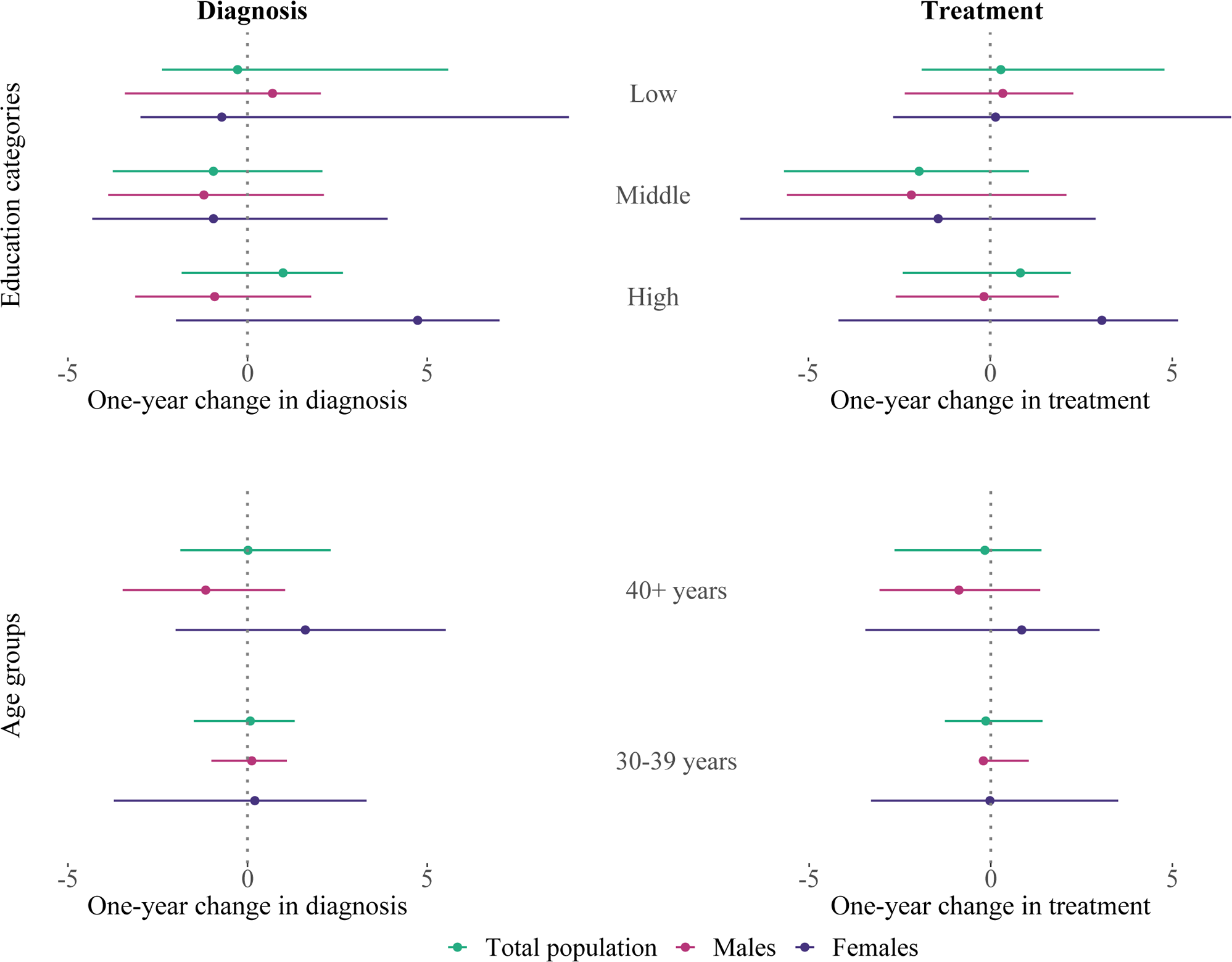
Regression Discontinuity Design results by education categories and age groups Note: The figure displays the effect estimates and 95% confidence intervals of home-based hypertension screening on confirmatory diagnosis (left panels) and drug treatment (right panels) by education category (top panels) and age group (bottom panels). Education categories are low education (up to primary school completed), middle education (secondary school completed), and high education (high school completed or higher). The model included linear local trends and triangular kernel weights, and estimated the effect within the mean squared error optimal bandwidth. The lower confidence interval of the treatment outcome of men aged 30-39 years was tiny and thus is not visible.

Lastly, we also do not find evidence of differential effects by a prior diagnosis of diabetes, heart attack, or stroke or city of residence (Chennai, Delhi) (Tables S9 to S12). While some differences in the direction of the effect can be consistently observed across groups, all point estimates were not statistically significant.

### 3.6 Sensitivity Analyses and Robustness

Our results were not sensitive to the choice of the MSE optimal bandwidth (Tables S13 and S14), the functional form used for the RDD estimation (Table S15), nor the recall period of the diagnosis and treatment questions (Table S16). Our results were consistent across the baseline-follow-up wave pairs (Tables S17 and S18). We also did not find evidence of heaping of BP values just above or below the threshold indicative of deliberate sorting around the threshold (Figure S3).

## 4 Discussion

We find that screening individuals’ BP in their homes and providing health information and a referral to those with elevated BP did not improve hypertension diagnosis or treatment rates or BP levels among adults living in two major Indian cities. Home-based screening was ineffective even when splitting the population by sex, age, education, prior diagnosis of cardiometabolic diseases, and city of residence. Our results reveal that lack of knowledge of one’s hypertension status and the need for hypertension care is likely not the primary reason behind low diagnosis and treatment rates in India. Rather, other structural and behavioral barriers may be responsible for the high levels of poor BP control. This is an especially important consideration for India as home- and community-based screening is being widely implemented as a strategy for improving hypertension control. ^7–9^

Our results contribute to a growing body of papers showing mixed effects of home-based screening interventions where individuals are provided health information and referred for further care. For example, in China, community-based hypertension screening reduced systolic BP among older adults. ^15^ A similar study in South Africa found that a home-based screening intervention reduced systolic BP only for females and younger males. ^16^ In Malawi, receiving a referral letter after home-based screening reduced BP and the probability of being hypertensive. ^14^ Lastly, in Germany, home-based screening and referral for hypertension had no long-term effect on cardiometabolic morbidity or mortality. ^27^ These mixed effects across contexts suggest that contextual circumstances substantially influence whether home-based screening interventions are effective. For example, intervention effectiveness may depend on (1) the underlying population and their beliefs and health-seeking behavior patterns; (2) the healthcare system in each context and how accessible care is; and (3) the design of the health information provided as part of the intervention including who provides individuals information and what information is provided. Identifying in which circumstances home-based screening interventions are effective and, additionally, how to design such interventions for maximum impact will be critical for future research and policy.

In our specific study context in urban India, there are several potential reasons why the home-based screening and information provision intervention did not increase hypertension diagnosis and treatment or reduce BP levels. At the individual level, the health information and referral provided by the survey team need to encourage individuals to visit a healthcare facility for a formal hypertension diagnosis, obtain a prescription, and then take the medication regularly. This requires that they understand the information and perceive hypertension to be a threat to their health. As hypertension is usually asymptomatic, individuals may not perceive it to be a disease that requires immediate and sustained action, both in terms of initiating and adhering to care. ^28–30^ Related to this is that hypertension care seeking may have low salience and be de-prioritized relative to other more pressing concerns. ^31^ A second barrier is related to direct and opportunity costs. In India, the majority of individuals seek hypertension care in private facilities, which requires them to pay for the services provided and the medication. ^32^ Furthermore, depending on the distance to the nearest facility, they might incur travel costs and lost income. ^33^ Another reason for not seeking treatment might be the fear of potential side effects or dependence on a drug or a preference for alternative treatments. ^28,29,33^

At the health system level, expanding hypertension screening coverage is essential for increasing hypertension diagnosis. Despite the Indian government’s NP-NCD framework mandate to opportunistically screen all adults aged 30 years or older, in reality, clinicians may not always screen patients for hypertension. ^10^ Thus, if hypertension is not perceived as a priority by healthcare workers and patients do not feel comfortable asking for a BP measurement, individuals following the referral by the home-based screening team might not obtain hypertension care although they acted on the health information received. ^29,33^

It is also uncertain whether clinicians provide correct follow-up care and treatment for those diagnosed as hypertensive and whether treatment is affordable and available.

The design of the home-based screening intervention may also have contributed to the null findings. The field team used non-standardized health messages that might have been conveyed differently by different field staff and their content might have depended on an individual’s BP level. These messages might not have been phrased in a way that adequately conveys the importance of hypertension control to the participant. Testing messages designed specifically for behavior change may improve the effectiveness of home-based screening. For example, designing health messages that cater to different levels of participant health literacy may improve the overall effectiveness of health messages. ^34,35^ Participants may react differently to loss-framed messages, i.e., emphasizing the risks of uncontrolled hypertension, or gain-framed messages, i.e., emphasizing the benefits of hypertension control. ^36^ Messages that directly address participant concerns around treatment side-effects and dependency or beliefs about symptoms are care seeking may also be more effective than traditional risk communication approaches. Evaluating the comparative performance of these different messaging approaches is an important next step.

This study has several limitations. First, our results are from two mega cities in India, and thus, the results cannot be generalized to the entire Indian population. Second, the RDD estimates the local average treatment effect, which is the effect for individuals with a standardized BP right at the threshold (i.e. zero). Home-based screening might affect health-seeking behavior differently for individuals with higher and more severe BP, but our results do not capture these potential effects. Third, the BP reading result at the health care facility might differ from the measurement recorded during the home-based screening. If the BP level was high during the home-based screening but normal at the facility, an individual would not have been diagnosed despite acting on the referral by the survey staff, resulting in an underestimation of the treatment effect. Fourth, we do not have any information on implementation fidelity. While one of the robustness checks (Figure S3) indicated that interviewers did not manipulate the measurement results to influence whether participants received the intervention or not, it was not possible to verify whether eligible participants received health information and referrals from the survey staff. Thus, the estimates should be interpreted as an intention-to-treat effect. ^16^ Furthermore, we have no information on how exactly the health messages were conveyed, which might have influenced whether a participant sought follow-up care.

Overall, our results suggest that home-based screening interventions that attempt to make individuals aware of their BP status and encourage them to seek further care may not be effective if other barriers to care-seeking are not also addressed. Exploring alternative communication approaches to address belief gaps and opportunity costs may help to improve the effectiveness of home-based screening in India and other contexts.

## Data Availability

Data are not publicly available but can be shared after a reasonable request to the data owners (Centre for Cardiometabolic Risk Reduction in South Asia (CARRS) team)

## 5 Ethical approval

This study uses secondary data and, thus, is designated as Non-Human Subjects Research not requiring ethical approval.

## 8 Author contributions

MT and SM accessed and verified the data, conducted the analysis, and wrote the first draft of this manuscript. All other authors critically revised the manuscript and approved its final version. All authors had access to the data and accepted the responsibility for publication. The corresponding author attests that all listed authors meet authorship criteria and that no others meeting the criteria have been omitted.

## 9 Supplementary material for the manuscript: Does home-based screening and health information provision improve hypertension diagnosis, treatment, and control? A regression discontinuity analysis in urban India

**Table S1:**
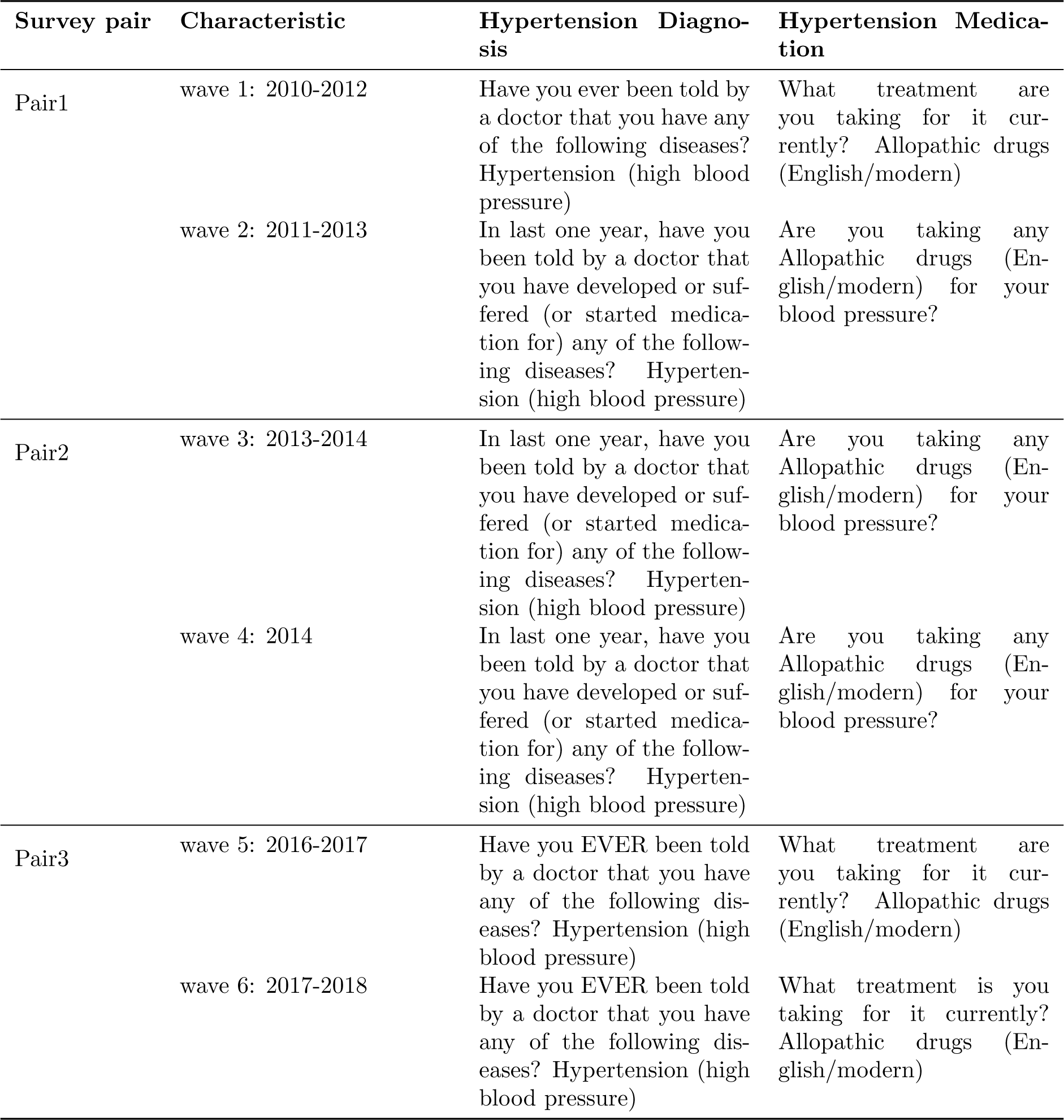
Survey Questions on hypertension diagnosis and treatment.

**Table S2:**
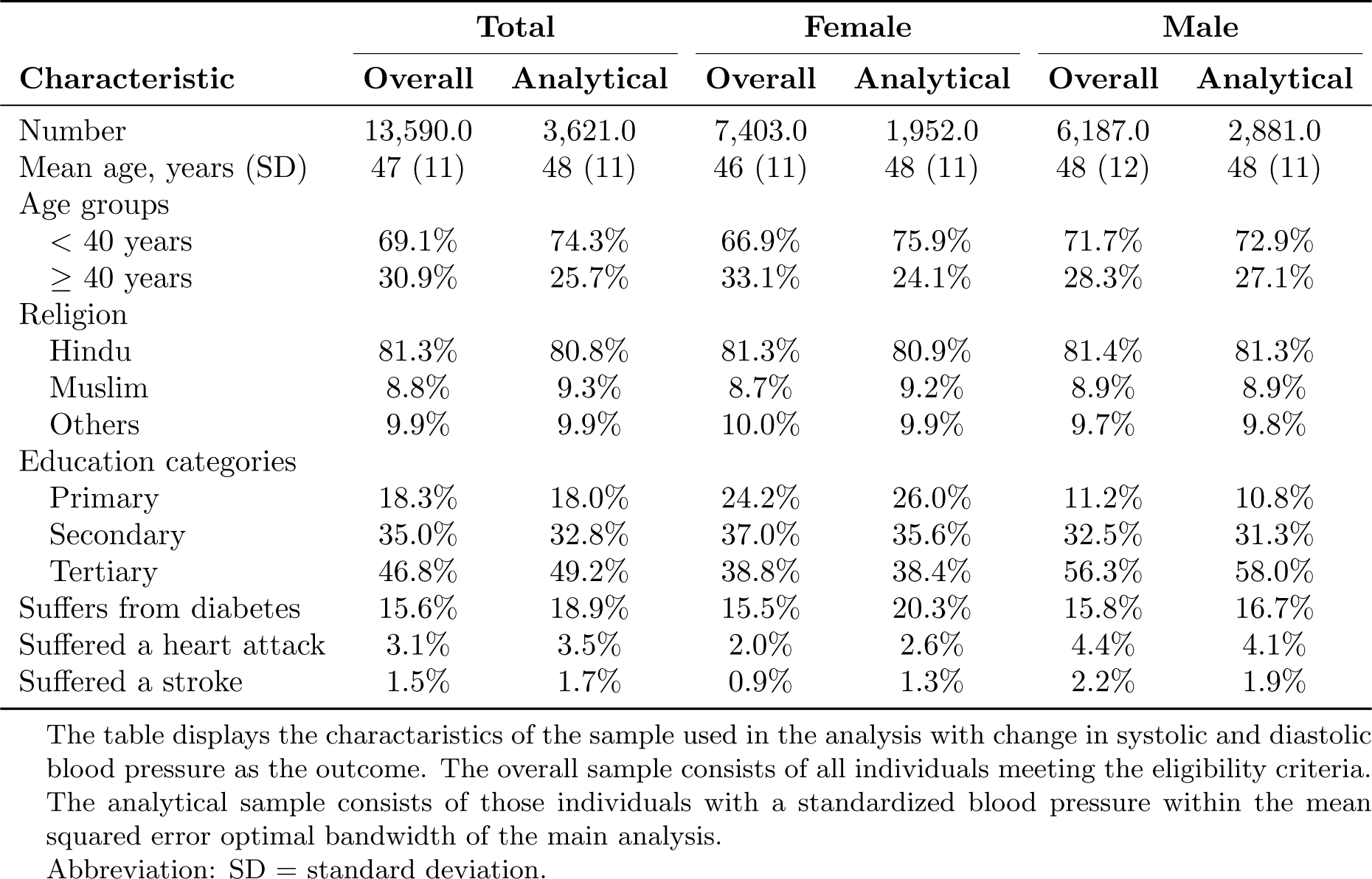
Characteristics of the overall and analytic samples for adults ages 30 and older, systolic and diastolic blood pressure.

| Characteristic | Total |  | Female |  | Male |  |
| --- | --- | --- | --- | --- | --- | --- |
|  | Overall | Analytical | Overall | Analytical | Overall | Analytical |
| Number | 13,590.0 | 3,621.0 | 7,403.0 | 1,952.0 | 6,187.0 | 2,881.0 |
| Mean age, years (SD) | 47 (11) | 48 (11) | 46 (11) | 48 (11) | 48 (12) | 48 (11) |
| Age groups |  |  |  |  |  |  |
| < 40 years | 69.1% | 74.3% | 66.9% | 75.9% | 71.7% | 72.9% |
| ≥ 40 years | 30.9% | 25.7% | 33.1% | 24.1% | 28.3% | 27.1% |
| Religion |  |  |  |  |  |  |
| Hindu | 81.3% | 80.8% | 81.3% | 80.9% | 81.4% | 81.3% |
| Muslim | 8.8% | 9.3% | 8.7% | 9.2% | 8.9% | 8.9% |
| Others | 9.9% | 9.9% | 10.0% | 9.9% | 9.7% | 9.8% |
| Education categories |  |  |  |  |  |  |
| Primary | 18.3% | 18.0% | 24.2% | 26.0% | 11.2% | 10.8% |
| Secondary | 35.0% | 32.8% | 37.0% | 35.6% | 32.5% | 31.3% |
| Tertiary | 46.8% | 49.2% | 38.8% | 38.4% | 56.3% | 58.0% |
| Suffers from diabetes | 15.6% | 18.9% | 15.5% | 20.3% | 15.8% | 16.7% |
| Suffered a heart attack | 3.1% | 3.5% | 2.0% | 2.6% | 4.4% | 4.1% |
| Suffered a stroke | 1.5% | 1.7% | 0.9% | 1.3% | 2.2% | 1.9% |

The table displays the characteristics of the sample used in the analysis with change in systolic and diastolic blood pressure as the outcome. The overall sample consists of all individuals meeting the eligibility criteria. The analytical sample consists of those individuals with a standardized blood pressure within the mean squared error optimal bandwidth of the main analysis.
Abbreviation: SD = standard deviation.

**Table S3:** Effect of home-based screening on hypertension diagnosis and treatment.

|  | Diagnosis | Treatment |
| --- | --- | --- |
| **Panel A: Total** |  |  |
| **Estimation results** |  |  |
| Coefficient | 0.12 | -0.16 |
| p-value | 0.82 | 0.49 |
| 95% CI | (-1.39 - 1.75) | (-2.18 - 1.03) |
| **Bandwidth details** |  |  |
| Bandwidth | 0.86 | 0.78 |
| Obs. within bandwidth | 7977 | 8566 |
| **Panel B: Female** |  |  |
| **Estimation results** |  |  |
| Coefficient | 1.55 | 0.64 |
| p-value | 0.40 | 0.84 |
| 95% CI | (-1.73 - 4.36) | (-3.15 - 2.56) |
| **Bandwidth details** |  |  |
| Bandwidth | 0.80 | 0.83 |
| Obs. within bandwidth | 3155 | 4216 |
| **Panel C: Male** |  |  |
| **Estimation results** |  |  |
| Coefficient | -0.82 | -0.76 |
| p-value | 0.32 | 0.37 |
| 95% CI | (-2.50 - 0.81) | (-2.28 - 0.86) |
| **Bandwidth details** |  |  |
| Bandwidth | 0.88 | 0.88 |
| Obs. within bandwidth | 4430 | 4884 |

Note: The model estimation included a local linear trend, triangular Kernel weights, and a data-driven bandwidth selection (mean squared error optimal bandwidth). The table displays the local average treatment effect in percentage points for individuals with a standardized blood pressure of zero and confidence intervals resulting from robust bias-corrected standard errors clustered at the individual-level.
Abbreviation: CI = confidence interval

**Table S4:** Effect of home-based screening on systolic and diastolic blood pressure.

|  | Systolic | Diastolic |
| --- | --- | --- |
| **Panel A: Total** |  |  |
| **Estimation results** |  |  |
| Coefficient | -1.84 | 0.48 |
| p-value | 0.03 | 0.39 |
| 95% CI | (-7.30 - -0.42) | (-0.94 - 2.41) |
| **Bandwidth details** |  |  |
| Bandwidth | 0.42 | 0.62 |
| Obs. within bandwidth | 3621 | 5438 |
| **Panel B: Female** |  |  |
| **Estimation results** |  |  |
| Coefficient | -2.26 | 0.17 |
| p-value | 0.06 | 0.60 |
| 95% CI | (-10.39 - 0.30) | (-1.84 - 3.18) |
| **Bandwidth details** |  |  |
| Bandwidth | 0.46 | 0.63 |
| Obs. within bandwidth | 1952 | 2695 |
| **Panel C: Male** |  |  |
| **Estimation results** |  |  |
| Coefficient | -0.38 | 0.85 |
| p-value | 0.24 | 0.50 |
| 95% CI | (-5.37 - 1.32) | (-1.42 - 2.90) |
| **Bandwidth details** |  |  |
| Bandwidth | 0.65 | 0.68 |
| Obs. within bandwidth | 2844 | 2996 |

Note: The model estimation included a local linear trend, triangular Kernel weights, and a data-driven bandwidth selection (mean squared error optimal bandwidth). The table displays the local average Diastolic effect in percentage points for individuals with a standardized blood pressure of zero and confidence intervals resulting from robust bias-corrected standard errors clustered at the individual-level.
Abbreviation: CI = confidence interval

**Table S5:**
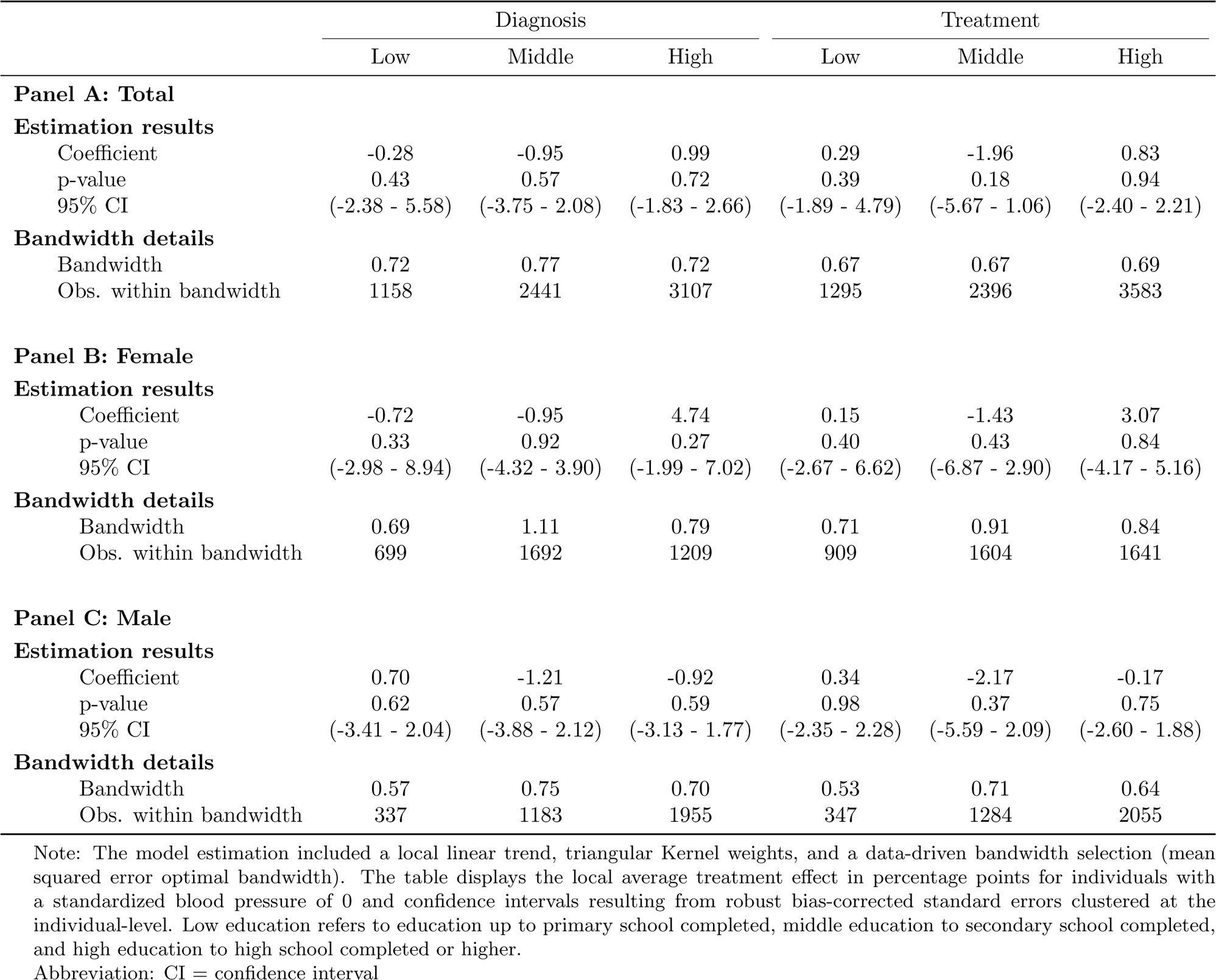
Effect of home-based screening by education categories.

**Table S6:** Effect of home-based screening by education categories.

|  | Systolic |  |  | Diastolic |  |  |
| --- | --- | --- | --- | --- | --- | --- |
|  | Low | Middle | High | Low | Middle | High |
| **Panel A: Total** |  |  |  |  |  |  |
| **Estimation results** |  |  |  |  |  |  |
| Coefficient | -2.35 | -3.80 | 0.73 | -0.08 | 0.35 | 0.80 |
| p-value | 0.12 | 0.01 | 0.94 | 0.91 | 0.99 | 0.23 |
| 95% CI | (-14.64 - 1.65) | (-12.83 - -2.03) | (-3.62 - 3.91) | (-4.16 - 3.72) | (-2.86 - 2.91) | (-0.93 - 3.82) |
| **Bandwidth details** |  |  |  |  |  |  |
| Bandwidth | 0.50 | 0.51 | 0.62 | 0.62 | 0.65 | 0.63 |
| Obs. within bandwidth | 839 | 1456 | 2586 | 1049 | 1933 | 2586 |
| **Panel B: Female** |  |  |  |  |  |  |
| **Estimation results** |  |  |  |  |  |  |
| Coefficient | -3.89 | -4.31 | 1.97 | -1.95 | 0.61 | 1.52 |
| p-value | 0.07 | 0.07 | 0.68 | 0.42 | 0.71 | 0.30 |
| 95% CI | (-18.29 - 0.56) | (-16.46 - 0.60) | (-5.11 - 7.88) | (-6.88 - 2.86) | (-3.09 - 4.57) | (-2.07 - 6.78) |
| **Bandwidth details** |  |  |  |  |  |  |
| Bandwidth | 0.49 | 0.50 | 0.74 | 0.53 | 0.69 | 0.60 |
| Obs. within bandwidth | 564 | 710 | 1169 | 613 | 1074 | 953 |
| **Panel C: Male** |  |  |  |  |  |  |
| **Estimation results** |  |  |  |  |  |  |
| Coefficient | 2.53 | -2.49 | 0.52 | 4.92 | 0.16 | 0.56 |
| p-value | 0.96 | 0.08 | 0.70 | 0.08 | 0.71 | 0.49 |
| 95% CI | (-12.00 - 12.63) | (-11.66 - 0.62) | (-4.72 - 3.17) | (-0.74 - 13.48) | (-4.83 - 3.30) | (-1.90 - 3.97) |
| **Bandwidth details** |  |  |  |  |  |  |
| Bandwidth | 0.69 | 0.67 | 0.72 | 0.68 | 0.66 | 0.59 |
| Obs. within bandwidth | 336 | 944 | 1826 | 325 | 930 | 1545 |

Note: The model estimation included a local linear trend, triangular Kernel weights, and a data-driven bandwidth selection (mean squared error optimal bandwidth). The table displays the local average treatment effect in percentage points for individuals with a standardized blood pressure of 0 and confidence intervals resulting from robust bias-corrected standard errors clustered at the individual-level. Low education refers to education up to primary school completed, middle education to secondary school completed, and high education to high school completed or higher.
Abbreviation: CI = confidence interval

**Table S7:** Effect of home-based screening by age groups.

|  | Diagnosis |  | Treatment |  |
| --- | --- | --- | --- | --- |
|  | 30-39 years | 40+ years | 30-39 years | 40+ years |
| **Panel A: Total** |  |  |  |  |
| **Estimation results** |  |  |  |  |
| Coefficient | 0.08 | 0.01 | -0.14 | -0.16 |
| p-value | 0.90 | 0.84 | 0.91 | 0.54 |
| 95% CI | (-1.49 - 1.31) | (-1.87 - 2.31) | (-1.26 - 1.43) | (-2.65 - 1.39) |
| **Bandwidth details** |  |  |  |  |
| Bandwidth | 0.60 | 0.85 | 0.66 | 0.83 |
| Obs. within bandwidth | 1444 | 5668 | 1752 | 6640 |
| **Panel B: Female** |  |  |  |  |
| **Estimation results** |  |  |  |  |
| Coefficient | 0.20 | 1.61 | -0.02 | 0.85 |
| p-value | 0.91 | 0.36 | 0.95 | 0.89 |
| 95% CI | (-3.72 - 3.31) | (-2.00 - 5.52) | (-3.30 - 3.51) | (-3.45 - 2.99) |
| **Bandwidth details** |  |  |  |  |
| Bandwidth | 0.69 | 0.81 | 0.69 | 0.98 |
| Obs. within bandwidth | 676 | 2405 | 730 | 3700 |
| **Panel C: Male** |  |  |  |  |
| **Estimation results** |  |  |  |  |
| Coefficient | 0.12 | -1.17 | -0.20 | -0.88 |
| p-value | 0.94 | 0.29 | 0.29 | 0.45 |
| 95% CI | (-1.01 - 1.09) | (-3.48 - 1.04) | (-0.31 - 1.04) | (-3.06 - 1.36) |
| **Bandwidth details** |  |  |  |  |
| Bandwidth | 0.57 | 0.94 | 0.61 | 0.85 |
| Obs. within bandwidth | 810 | 3373 | 901 | 3543 |

Note: The model estimation included a local linear trend, triangular Kernel weights, and a data-driven bandwidth selection (mean squared error optimal bandwidth). The table displays the local average treatment effect in percentage points for individuals with a standardized blood pressure of zero and confidence intervals resulting from robust bias-corrected standard errors clustered at the individual-level.
Abbreviation: CI = confidence interval

**Table S8:** Effect of home-based screening by age groups.

|  | Systolic |  | Diastolic |  |
| --- | --- | --- | --- | --- |
|  | 30-39 years | 40+ years | 30-39 years | 40+ years |
| **Panel A: Total** |  |  |  |  |
| **Estimation results** |  |  |  |  |
| Coefficient | 0.02 | -2.39 | -0.09 | 0.81 |
| p-value | 0.78 | 0.05 | 0.81 | 0.37 |
| 95% CI | (-5.56 - 4.15) | (-8.18 - -0.00) | (-4.21 - 3.29) | (-0.98 - 2.64) |
| **Bandwidth details** |  |  |  |  |
| Bandwidth | 0.72 | 0.39 | 0.61 | 0.67 |
| Obs. within bandwidth | 1464 | 2692 | 1294 | 4505 |
| **Panel B: Female** |  |  |  |  |
| **Estimation results** |  |  |  |  |
| Coefficient | -1.00 | -2.86 | -0.93 | 0.63 |
| p-value | 0.88 | 0.05 | 0.71 | 0.75 |
| 95% CI | (-7.63 - 6.54) | (-12.65 - -0.13) | (-4.87 - 7.13) | (-2.37 - 3.30) |
| **Bandwidth details** |  |  |  |  |
| Bandwidth | 0.95 | 0.43 | 0.70 | 0.59 |
| Obs. within bandwidth | 870 | 1367 | 616 | 1920 |
| **Panel C: Male** |  |  |  |  |
| **Estimation results** |  |  |  |  |
| Coefficient | 0.60 | -0.93 | 1.07 | 0.88 |
| p-value | 0.78 | 0.29 | 0.69 | 0.33 |
| 95% CI | (-5.91 - 4.44) | (-6.53 - 1.98) | (-4.50 - 2.97) | (-1.17 - 3.52) |
| **Bandwidth details** |  |  |  |  |
| Bandwidth | 0.79 | 0.58 | 0.77 | 0.77 |
| Obs. within bandwidth | 844 | 1955 | 841 | 2458 |

Note: The model estimation included a local linear trend, triangular Kernel weights, and a data-driven bandwidth selection (mean squared error optimal bandwidth). The table displays the local average treatment effect in percentage points for individuals with a standardized blood pressure of zero and confidence intervals resulting from robust bias-corrected standard errors clustered at the individual-level.
Abbreviation: CI = confidence interval

**Table S9:**
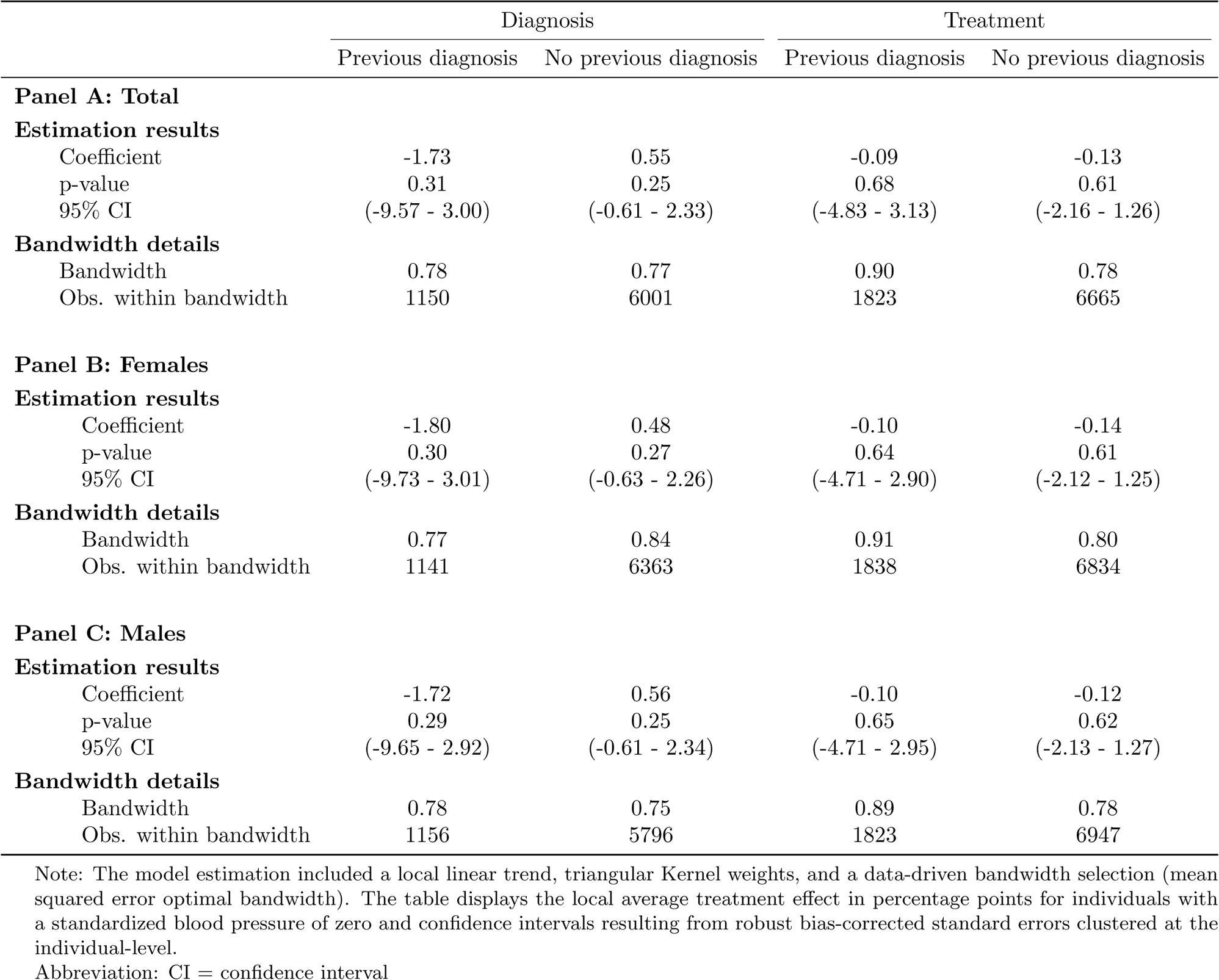
Effect of home-based screening by previous diabetes, heart attack, or stroke diagnosis.

|  | Diagnosis |  | Treatment |  |
| --- | --- | --- | --- | --- |
|  | Previous diagnosis | No previous diagnosis | Previous diagnosis | No previous diagnosis |
| **Panel A: Total** |  |  |  |  |
| **Estimation results** |  |  |  |  |
| Coefficient | -1.73 | 0.55 | -0.09 | -0.13 |
| p-value | 0.31 | 0.25 | 0.68 | 0.61 |
| 95% CI | (-9.57 - 3.00) | (-0.61 - 2.33) | (-4.83 - 3.13) | (-2.16 - 1.26) |
| **Bandwidth details** |  |  |  |  |
| Bandwidth | 0.78 | 0.77 | 0.90 | 0.78 |
| Obs. within bandwidth | 1150 | 6001 | 1823 | 6665 |
| **Panel B: Females** |  |  |  |  |
| **Estimation results** |  |  |  |  |
| Coefficient | -1.80 | 0.48 | -0.10 | -0.14 |
| p-value | 0.30 | 0.27 | 0.64 | 0.61 |
| 95% CI | (-9.73 - 3.01) | (-0.63 - 2.26) | (-4.71 - 2.90) | (-2.12 - 1.25) |
| **Bandwidth details** |  |  |  |  |
| Bandwidth | 0.77 | 0.84 | 0.91 | 0.80 |
| Obs. within bandwidth | 1141 | 6363 | 1838 | 6834 |
| **Panel C: Males** |  |  |  |  |
| **Estimation results** |  |  |  |  |
| Coefficient | -1.72 | 0.56 | -0.10 | -0.12 |
| p-value | 0.29 | 0.25 | 0.65 | 0.62 |
| 95% CI | (-9.65 - 2.92) | (-0.61 - 2.34) | (-4.71 - 2.95) | (-2.13 - 1.27) |
| **Bandwidth details** |  |  |  |  |
| Bandwidth | 0.78 | 0.75 | 0.89 | 0.78 |
| Obs. within bandwidth | 1156 | 5796 | 1823 | 6947 |

Note: The model estimation included a local linear trend, triangular Kernel weights, and a data-driven bandwidth selection (mean squared error optimal bandwidth). The table displays the local average treatment effect in percentage points for individuals with a standardized blood pressure of zero and confidence intervals resulting from robust bias-corrected standard errors clustered at the individual-level.
Abbreviation: CI = confidence interval

**Table S10:**
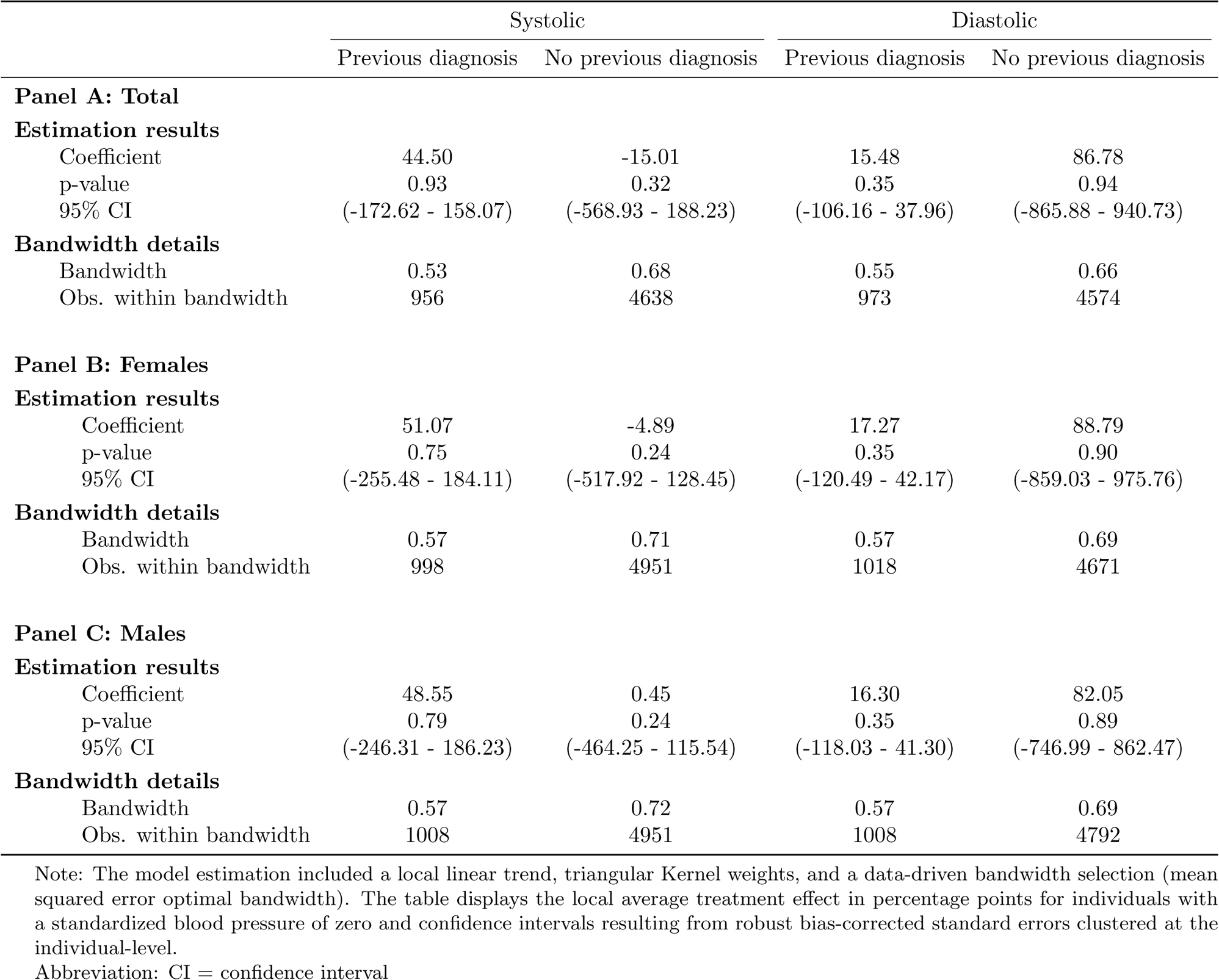
Effect of home-based screening by previous diabetes, heart attack, or stroke diagnosis.

|  | Systolic |  | Diastolic |  |
| --- | --- | --- | --- | --- |
|  | Previous diagnosis | No previous diagnosis | Previous diagnosis | No previous diagnosis |
| **Panel A: Total** |  |  |  |  |
| **Estimation results** |  |  |  |  |
| Coefficient | 44.50 | -15.01 | 15.48 | 86.78 |
| p-value | 0.93 | 0.32 | 0.35 | 0.94 |
| 95% CI | (-172.62 - 158.07) | (-568.93 - 188.23) | (-106.16 - 37.96) | (-865.88 - 940.73) |
| **Bandwidth details** |  |  |  |  |
| Bandwidth | 0.53 | 0.68 | 0.55 | 0.66 |
| Obs. within bandwidth | 956 | 4638 | 973 | 4574 |
| **Panel B: Females** |  |  |  |  |
| **Estimation results** |  |  |  |  |
| Coefficient | 51.07 | -4.89 | 17.27 | 88.79 |
| p-value | 0.75 | 0.24 | 0.35 | 0.90 |
| 95% CI | (-255.48 - 184.11) | (-517.92 - 128.45) | (-120.49 - 42.17) | (-859.03 - 975.76) |
| **Bandwidth details** |  |  |  |  |
| Bandwidth | 0.57 | 0.71 | 0.57 | 0.69 |
| Obs. within bandwidth | 998 | 4951 | 1018 | 4671 |
| **Panel C: Males** |  |  |  |  |
| **Estimation results** |  |  |  |  |
| Coefficient | 48.55 | 0.45 | 16.30 | 82.05 |
| p-value | 0.79 | 0.24 | 0.35 | 0.89 |
| 95% CI | (-246.31 - 186.23) | (-464.25 - 115.54) | (-118.03 - 41.30) | (-746.99 - 862.47) |
| **Bandwidth details** |  |  |  |  |
| Bandwidth | 0.57 | 0.72 | 0.57 | 0.69 |
| Obs. within bandwidth | 1008 | 4951 | 1008 | 4792 |

Note: The model estimation included a local linear trend, triangular Kernel weights, and a data-driven bandwidth selection (mean squared error optimal bandwidth). The table displays the local average treatment effect in percentage points for individuals with a standardized blood pressure of zero and confidence intervals resulting from robust bias-corrected standard errors clustered at the individual-level.
Abbreviation: CI = confidence interval

**Table S11:** Effect of home-based screening by city.

|  | Diagnosis |  | Treatment |  |
| --- | --- | --- | --- | --- |
|  | Chennai | Delhi | Chennai | Delhi |
| **Panel A: Total** |  |  |  |  |
| **Estimation results** |  |  |  |  |
| Coefficient | 1.29 | -0.90 | 0.68 | -0.83 |
| p-value | 0.33 | 0.30 | 0.73 | 0.19 |
| 95% CI | (-1.32 - 3.96) | (-2.85 - 0.87) | (-2.13 - 3.03) | (-3.05 - 0.61) |
| **Bandwidth details** |  |  |  |  |
| Bandwidth | 0.86 | 0.97 | 0.90 | 0.76 |
| Obs. within bandwidth | 3827 | 4529 | 4525 | 4346 |
| **Panel B: Females** |  |  |  |  |
| **Estimation results** |  |  |  |  |
| Coefficient | 1.28 | -0.90 | 0.63 | -0.83 |
| p-value | 0.36 | 0.30 | 0.75 | 0.20 |
| 95% CI | (-1.41 - 3.88) | (-2.85 - 0.87) | (-2.15 - 2.97) | (-3.01 - 0.62) |
| **Bandwidth details** |  |  |  |  |
| Bandwidth | 0.87 | 0.96 | 0.91 | 0.77 |
| Obs. within bandwidth | 3764 | 4445 | 4538 | 4317 |
| **Panel C: Males** |  |  |  |  |
| **Estimation results** |  |  |  |  |
| Coefficient | 1.28 | -0.91 | 0.66 | -0.82 |
| p-value | 0.33 | 0.32 | 0.74 | 0.20 |
| 95% CI | (-1.34 - 3.94) | (-2.82 - 0.92) | (-2.14 - 3.00) | (-3.01 - 0.64) |
| **Bandwidth details** |  |  |  |  |
| Bandwidth | 0.86 | 0.94 | 0.89 | 0.77 |
| Obs. within bandwidth | 3807 | 4360 | 4571 | 4317 |

Note: The model estimation included a local linear trend, triangular Kernel weights, and a data-driven bandwidth selection (mean squared error optimal bandwidth). The table displays the local average treatment effect in percentage points for individuals with a standardized blood pressure of zero and confidence intervals resulting from robust bias-corrected standard errors clustered at the individual-level.
Abbreviation: CI = confidence interval

**Table S12:**
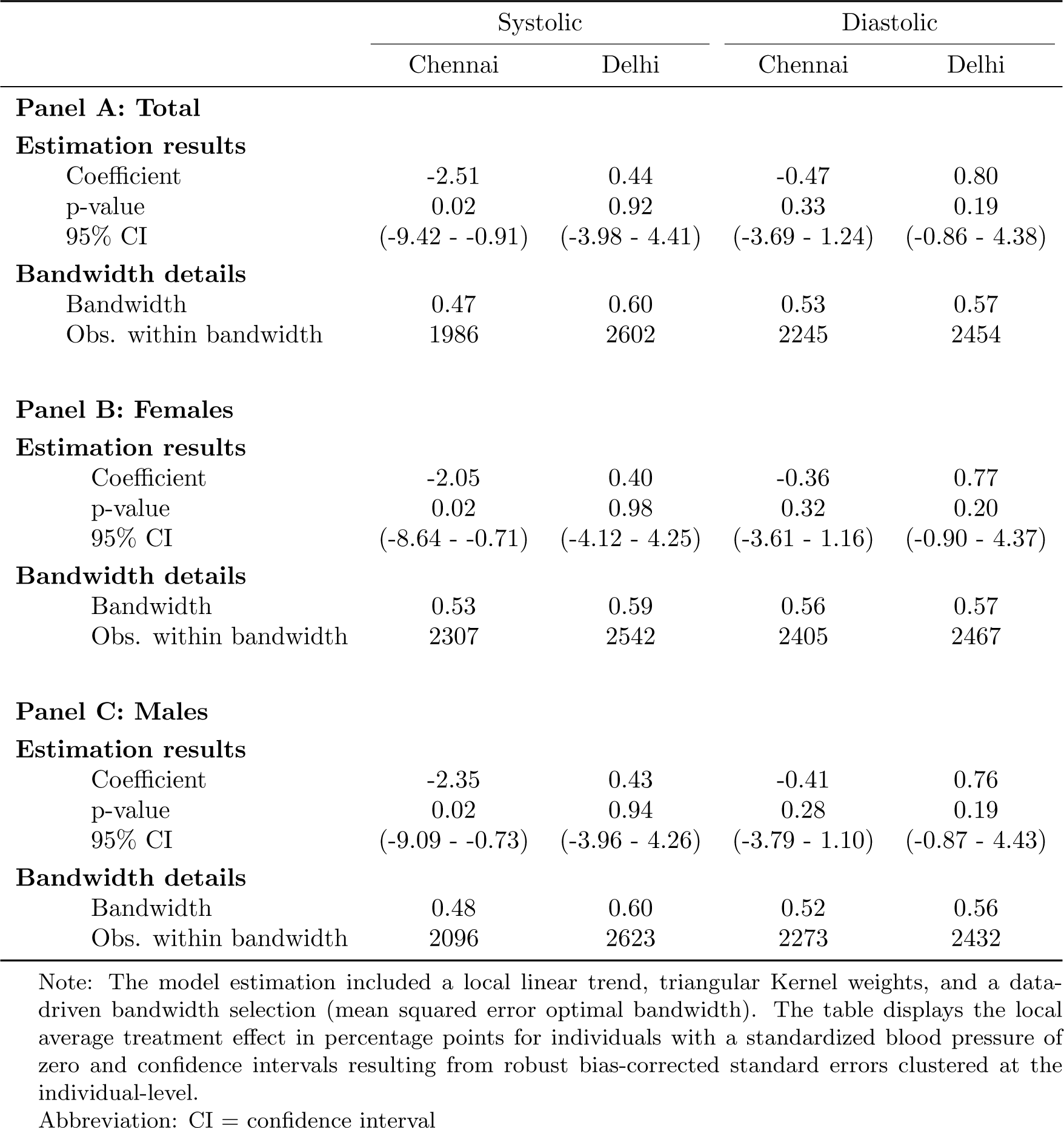
Effect of home-based screening by city.

**Table S13:** Effect of home-based screening on hypertension diagnosis with varying bandwidths.

|  | (1) | (2) | (3) | (4) | (5) | (6) | (7) | (8) | (9) |
| --- | --- | --- | --- | --- | --- | --- | --- | --- | --- |
| **Panel A: Total** |  |  |  |  |  |  |  |  |  |
| **Estimation results** |  |  |  |  |  |  |  |  |  |
| Coefficient | 0.12 | 0.13 | 0.17 | 0.16 | 0.12 | 0.09 | 0.09 | 0.09 | 0.1 |
| p-value | 0.7 | 0.79 | 0.87 | 0.87 | 0.82 | 0.79 | 0.81 | 0.84 | 0.86 |
| 95% CI | (-1.39 - 2.06) | (-1.45 - 1.91) | (-1.51 - 1.77) | (-1.47 - 1.73) | (-1.39 - 1.75) | (-1.33 - 1.75) | (-1.32 - 1.7) | (-1.33 - 1.63) | (-1.33 - 1.59) |
| **Bandwidth details** |  |  |  |  |  |  |  |  |  |
| Bandwidth | 0.66 | 0.71 | 0.76 | 0.81 | 0.86 | 0.91 | 0.96 | 1.01 | 1.06 |
| Obs. within bandwidth | 5915 | 6398 | 7132 | 7611 | 7977 | 8386 | 8786 | 9217 | 9528 |
| **Panel B: Female** |  |  |  |  |  |  |  |  |  |
| **Estimation results** |  |  |  |  |  |  |  |  |  |
| Coefficient | 1.42 | 1.41 | 1.45 | 1.51 | 1.55 | 1.5 | 1.41 | 1.32 | 1.3 |
| p-value | 0.34 | 0.33 | 0.37 | 0.39 | 0.4 | 0.35 | 0.29 | 0.25 | 0.25 |
| 95% CI | (-1.84 - 5.32) | (-1.72 - 5.07) | (-1.76 - 4.76) | (-1.77 - 4.52) | (-1.73 - 4.36) | (-1.55 - 4.37) | (-1.32 - 4.47) | (-1.16 - 4.51) | (-1.13 - 4.43) |
| **Bandwidth details** |  |  |  |  |  |  |  |  |  |
| Bandwidth | 0.6 | 0.65 | 0.7 | 0.75 | 0.8 | 0.85 | 0.9 | 0.95 | 1 |
| Obs. within bandwidth | 2316 | 2523 | 2742 | 2972 | 3155 | 3537 | 3762 | 3973 | 4134 |
| **Panel C: Male** |  |  |  |  |  |  |  |  |  |
| **Estimation results** |  |  |  |  |  |  |  |  |  |
| Coefficient | -0.86 | -0.86 | -0.84 | -0.84 | -0.82 | -0.79 | -0.74 | -0.7 | -0.67 |
| p-value | 0.47 | 0.42 | 0.37 | 0.35 | 0.32 | 0.28 | 0.24 | 0.22 | 0.2 |
| 95% CI | (-2.36 - 1.09) | (-2.43 - 1.01) | (-2.49 - 0.93) | (-2.49 - 0.87) | (-2.5 - 0.81) | (-2.52 - 0.74) | (-2.55 - 0.65) | (-2.56 - 0.59) | (-2.55 - 0.54) |
| **Bandwidth details** |  |  |  |  |  |  |  |  |  |
| Bandwidth | 0.68 | 0.73 | 0.78 | 0.83 | 0.88 | 0.93 | 0.98 | 1.03 | 1.08 |
| Obs. within bandwidth | 3421 | 3672 | 3836 | 4211 | 4430 | 4611 | 4769 | 4936 | 5133 |

The model estimation included a local linear trend, triangular Kernel weights, and a data-driven bandwidth selection (mean squared error optimal bandwidth). The table displays the local average treatment effect in percentage points for individuals with a standardized blood pressure of zero and CIs resulting from robust bias-corrected standard errors clustered at the individual-level. The bandwidth was varied by +/- 0.2 in intervals of 0.05, column (5) contains the results of the main analysis. Abbreviation: CI = confidence interval

**Table S14:** Effect of home-based screening on hypertension treatment with varying bandwidths.

|  | (1) | (2) | (3) | (4) | (5) | (6) | (7) | (8) | (9) |
| --- | --- | --- | --- | --- | --- | --- | --- | --- | --- |
| **Panel A: Total** |  |  |  |  |  |  |  |  |  |
| **Estimation results** |  |  |  |  |  |  |  |  |  |
| Coefficient | -0.37 | -0.3 | -0.24 | -0.17 | -0.16 | -0.15 | -0.14 | -0.11 | -0.09 |
| p-value | 0.61 | 0.53 | 0.48 | 0.44 | 0.49 | 0.53 | 0.55 | 0.55 | 0.56 |
| 95% CI | (-2.32 - 1.37) | (-2.35 - 1.2) | (-2.33 - 1.09) | (-2.3 - 1.01) | (-2.18 - 1.03) | (-2.06 - 1.06) | (-1.98 - 1.06) | (-1.93 - 1.03) | (-1.88 - 1.02) |
| **Bandwidth details** |  |  |  |  |  |  |  |  |  |
| Bandwidth | 0.58 | 0.63 | 0.68 | 0.73 | 0.78 | 0.83 | 0.88 | 0.93 | 0.98 |
| Obs. within bandwidth | 6212 | 6695 | 7223 | 7745 | 8566 | 9075 | 9530 | 9985 | 10451 |
| **Panel B: Female** |  |  |  |  |  |  |  |  |  |
| **Estimation results** |  |  |  |  |  |  |  |  |  |
| Coefficient | 0.2 | 0.38 | 0.52 | 0.61 | 0.64 | 0.66 | 0.72 | 0.76 | 0.81 |
| p-value | 0.85 | 0.76 | 0.74 | 0.76 | 0.84 | 0.91 | 0.93 | 0.96 | 1 |
| 95% CI | (-3.6 - 2.97) | (-3.65 - 2.67) | (-3.56 - 2.53) | (-3.4 - 2.49) | (-3.15 - 2.56) | (-2.94 - 2.62) | (-2.85 - 2.59) | (-2.72 - 2.59) | (-2.6 - 2.59) |
| **Bandwidth details** |  |  |  |  |  |  |  |  |  |
| Bandwidth | 0.63 | 0.68 | 0.73 | 0.78 | 0.83 | 0.88 | 0.93 | 0.98 | 1.03 |
| Obs. within bandwidth | 3008 | 3409 | 3677 | 3929 | 4216 | 4417 | 4631 | 4877 | 5129 |
| **Panel C: Male** |  |  |  |  |  |  |  |  |  |
| **Estimation results** |  |  |  |  |  |  |  |  |  |
| Coefficient | -0.75 | -0.73 | -0.75 | -0.76 | -0.76 | -0.76 | -0.74 | -0.74 | -0.71 |
| p-value | 0.43 | 0.39 | 0.4 | 0.39 | 0.37 | 0.36 | 0.33 | 0.31 | 0.28 |
| 95% CI | (-2.47 - 1.04) | (-2.45 - 0.95) | (-2.37 - 0.94) | (-2.32 - 0.9) | (-2.28 - 0.86) | (-2.25 - 0.81) | (-2.23 - 0.75) | (-2.2 - 0.71) | (-2.21 - 0.64) |
| **Bandwidth details** |  |  |  |  |  |  |  |  |  |
| Bandwidth | 0.68 | 0.73 | 0.78 | 0.83 | 0.88 | 0.93 | 0.98 | 1.03 | 1.08 |
| Obs. within bandwidth | 3968 | 4197 | 4460 | 4717 | 4884 | 5219 | 5404 | 5615 | 5804 |

The model estimation included a local linear trend, triangular Kernel weights, and a data-driven bandwidth selection (mean squared error optimal bandwidth). The table displays the local average treatment effect in percentage points for individuals with a standardized blood pressure of zero and CIs resulting from robust bias-corrected standard errors clustered at the individual-level. The bandwidth was varied by +/- 0.2 in intervals of 0.05, column (5) contains the results of the main analysis. Abbreviation: CI = confidence interval

**Table S15:** Effect of home-based screening on hypertension diagnosis and treatment: Local quadratic trend.

|  | Diagnosis | Treatment |
| --- | --- | --- |
| **Panel A: Total** |  |  |
| **Estimation results** |  |  |
| Coefficient | 0.13 | -0.26 |
| p-value | 0.68 | 0.47 |
| 95% CI | (-1.44 - 2.2) | (-2.25 - 1.05) |
| **Bandwidth details** |  |  |
| Bandwidth | 1.04 | 1.29 |
| Obs. within bandwidth | 9528 | 13225 |
| **Panel B: Female** |  |  |
| **Estimation results** |  |  |
| Coefficient | 1.63 | 0.24 |
| p-value | 0.4 | 0.72 |
| 95% CI | (-1.97 - 4.89) | (-3.7 - 2.57) |
| **Bandwidth details** |  |  |
| Bandwidth | 1.1 | 1.19 |
| Obs. within bandwidth | 4589 | 5973 |
| **Panel C: Male** |  |  |
| **Estimation results** |  |  |
| Coefficient | -0.81 | 0.27 |
| p-value | 0.93 | 0.06 |
| 95% CI | (-1.72 - 1.88) | (-0.07 - 2.78) |
| **Bandwidth details** |  |  |
| Bandwidth | 0.82 | 0.72 |
| Obs. within bandwidth | 4074 | 3623 |

The model estimation included a local quadratic trend, triangular Kernel weights, and a data-driven bandwidth selection (mean squared error optimal bandwidth). The table displays the local average treatment effect in percentage points for individuals with a standardized blood pressure of zero and CIs resulting from robust bias-corrected standard errors clustered at the individual-level. Abbreviation: CI = confidence interval

**Table S16:** Effect of home-based screening restricted to follow-up surveys conducted within the 12 month recall period.

|  | Diagnosis | Treatment |
| --- | --- | --- |
| **Panel A: Total** |  |  |
| **Estimation results** |  |  |
| Coefficient | -0.03 | -0.4 |
| p-value | 0.92 | 0.31 |
| 95% CI | (-1.46 - 1.63) | (-2.68 - 0.86) |
| **Bandwidth details** |  |  |
| Bandwidth | 1 | 0.96 |
| Obs. within bandwidth | 7283 | 8346 |
| **Panel B: Female** |  |  |
| **Estimation results** |  |  |
| Coefficient | 0.62 | 0.59 |
| p-value | 0.41 | 0.82 |
| 95% CI | (-1.79 - 4.37) | (-3.63 - 2.89) |
| **Bandwidth details** |  |  |
| Bandwidth | 1.01 | 0.95 |
| Obs. within bandwidth | 3359 | 3968 |
| **Panel C: Male** |  |  |
| **Estimation results** |  |  |
| Coefficient | -0.72 | -0.72 |
| p-value | 0.34 | 0.34 |
| 95% CI | (-1.81 - 0.63) | (-1.81 - 0.63) |
| **Bandwidth details** |  |  |
| Bandwidth | 0.66 | 0.66 |
| Obs. within bandwidth | 2645 | 2645 |

The model estimation included a local linear trend, triangular Kernel weights, and a data-driven bandwidth selection (mean squared error optimal bandwidth). The table displays the local average treatment effect in percentage points for individuals with a standardized blood pressure of zero and CIs resulting from robust bias-corrected standard errors clustered at the individual-level.
Abbreviation: CI = confidence interval

**Table S17:** Effect of home-based screening by survey pair.

|  | Diagnosis |  |  | Treatment |  |  |
| --- | --- | --- | --- | --- | --- | --- |
|  | Pair 1 | Pair 2 | Pair 3 | Pair 1 | Pair 2 | Pair 3 |
| **Panel A: Total** |  |  |  |  |  |  |
| **Estimation results** |  |  |  |  |  |  |
| Coefficient | 1.67 | -0.30 | -1.39 | 1.66 | -1.43 | -1.13 |
| p-value | 0.09 | 0.55 | 0.65 | 0.12 | 0.17 | 0.47 |
| 95% CI | (-0.33 - 4.15) | (-2.64 - 1.41) | (-5.62 - 3.49) | (-0.45 - 4.00) | (-5.15 - 0.89) | (-4.76 - 2.18) |
| **Bandwidth details** |  |  |  |  |  |  |
| Bandwidth | 0.76 | 0.79 | 0.66 | 0.67 | 0.64 | 0.87 |
| Obs. within bandwidth | 2562 | 2445 | 1658 | 2502 | 2518 | 2716 |
| **Panel B: Female** |  |  |  |  |  |  |
| **Estimation results** |  |  |  |  |  |  |
| Coefficient | 2.91 | 0.10 | 1.01 | 2.83 | -1.91 | 0.50 |
| p-value | 0.09 | 0.83 | 0.96 | 0.11 | 0.16 | 0.99 |
| 95% CI | (-0.52 - 7.91) | (-3.20 - 2.57) | (-8.03 - 8.42) | (-0.74 - 7.25) | (-8.55 - 1.40) | (-6.19 - 6.24) |
| **Bandwidth details** |  |  |  |  |  |  |
| Bandwidth | 0.80 | 0.74 | 0.68 | 0.79 | 0.66 | 0.87 |
| Obs. within bandwidth | 1248 | 1041 | 813 | 1382 | 1259 | 1325 |
| **Panel C: Male** |  |  |  |  |  |  |
| **Estimation results** |  |  |  |  |  |  |
| Coefficient | 0.46 | -0.75 | -3.18 | 0.60 | -0.84 | -2.30 |
| p-value | 0.56 | 0.63 | 0.21 | 0.75 | 0.55 | 0.24 |
| 95% CI | (-1.53 - 2.82) | (-3.43 - 2.06) | (-7.15 - 1.56) | (-1.87 - 2.62) | (-4.07 - 2.16) | (-6.03 - 1.48) |
| **Bandwidth details** |  |  |  |  |  |  |
| Bandwidth | 0.90 | 0.74 | 0.78 | 0.73 | 0.83 | 0.73 |
| Obs. within bandwidth | 1685 | 1250 | 1108 | 1412 | 1699 | 1219 |

The model estimation included a local linear trend, triangular Kernel weights, and a data-driven bandwidth selection (mean squared error optimal bandwidth). The table displays the local average treatment effect in percentage points for individuals with a standardized blood pressure of zero and CIs resulting from robust bias-corrected standard errors clustered at the individual-level. Abbreviation: CI = confidence interval

**Table S18:** Effect of home-based screening by survey pair on blood pressure.

|  | Systolic |  | Diastolic |  |
| --- | --- | --- | --- | --- |
|  | Pair A | Pair B | Pair A | Pair B |
| **Panel A: Total** |  |  |  |  |
| **Estimation results** |  |  |  |  |
| Coefficient | 1.22 | -8.02 | 0.93 | 0.10 |
| p-value | 0.31 | 0.00 | 0.16 | 0.97 |
| 95% CI | (-1.65 - 5.20) | (-17.13 - -4.64) | (-0.65 - 4.07) | (-2.01 - 1.92) |
| **Bandwidth details** |  |  |  |  |
| Bandwidth | 0.70 | 0.29 | 0.61 | 0.93 |
| Obs. within bandwidth | 3313 | 1128 | 2920 | 3598 |
| **Panel B: Female** |  |  |  |  |
| **Estimation results** |  |  |  |  |
| Coefficient | 0.14 | -7.59 | -0.01 | 0.35 |
| p-value | 0.80 | 0.03 | 0.63 | 0.88 |
| 95% CI | (-5.05 - 6.51) | (-16.89 - -0.81) | (-2.76 - 4.55) | (-2.94 - 3.42) |
| **Bandwidth details** |  |  |  |  |
| Bandwidth | 0.69 | 0.36 | 0.63 | 0.65 |
| Obs. within bandwidth | 1657 | 722 | 1477 | 1290 |
| **Panel C: Male** |  |  |  |  |
| **Estimation results** |  |  |  |  |
| Coefficient | 2.21 | -6.45 | 1.80 | -0.51 |
| p-value | 0.16 | 0.00 | 0.12 | 0.53 |
| 95% CI | (-1.08 - 6.73) | (-17.49 - -3.89) | (-0.61 - 5.06) | (-4.84 - 2.48) |
| **Bandwidth details** |  |  |  |  |
| Bandwidth | 0.78 | 0.42 | 0.68 | 0.57 |
| Obs. within bandwidth | 1828 | 851 | 1580 | 1156 |

Pair A: Wave 1 and 3 and Pair B: Wave 3 and 5. The model estimation included a local linear trend, triangular Kernel weights, and a data-driven bandwidth selection (mean squared error optimal bandwidth). The table displays the local average treatment effect for individuals with a standardized blood pressure of zero and CIs resulting from robust bias-corrected standard errors clustered at the individual-level.
Abbreviation: CI = confidence interval

**Figure S1:**
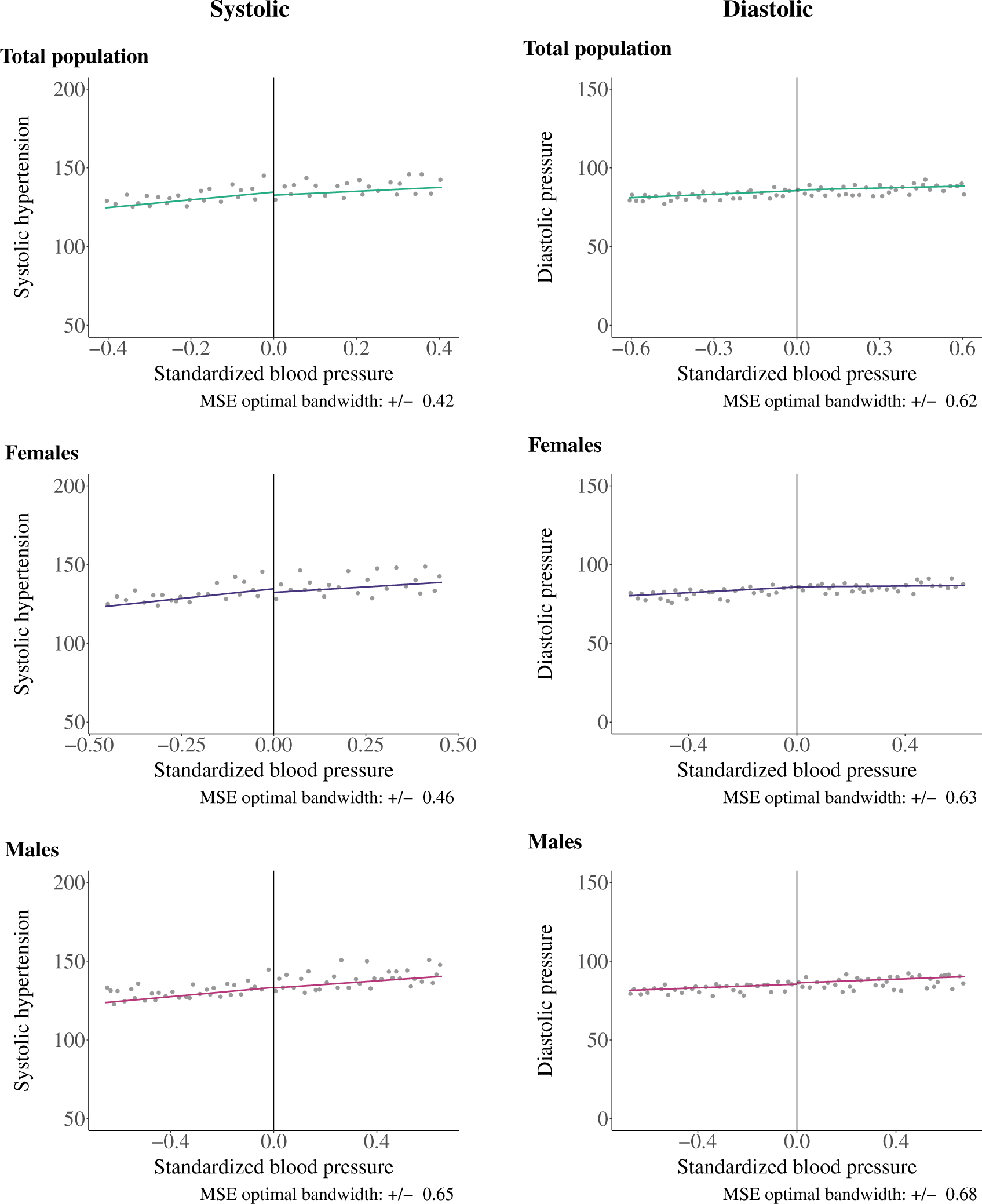
Systolic blood pressure (left panel) and diastolic blood pressure (right panel) among adults aged 30 and older Note: These plots only show observations within each mean squared error optimal bandwidth. The vertical line indicates the threshold set at a standardized BP of zero. The colored lines represent the linear trends fitted separately for individuals with a standardized BP below zero and those with a standardized BP of zero or higher. ^0^Note: The vertical line in the top panel marks the threshold at a systolic BP of 140 mmHg and in the bottom panel at a diastolic BP of 90 mmHg.

**Figure S2:**
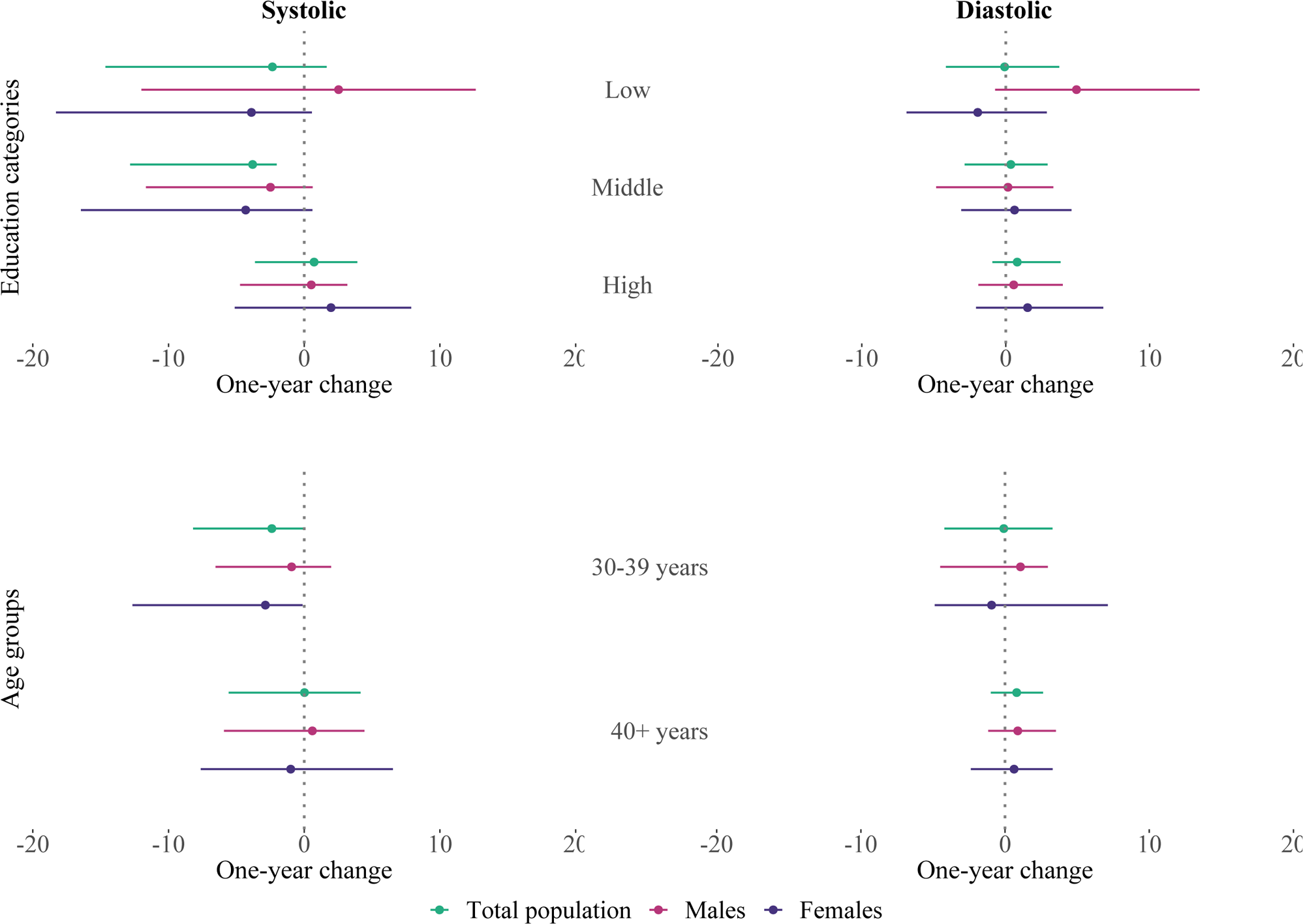
Regression Discontinuity Design results by education categories and age groups Note: The figure displays the effect estimates and 95% confidence intervals of home-based hypertension screening on systolic (left panels) and diastolic blood pressure (right panels) by education category (top panels) and age group (bottom panels). Education categories are low education (up to primary school completed), middle education (secondary school completed), and high education (high school completed or higher). The model included linear local trends and triangular kernel weights, and estimated the effect within the mean squared error optimal bandwidth.

**Figure S3:**
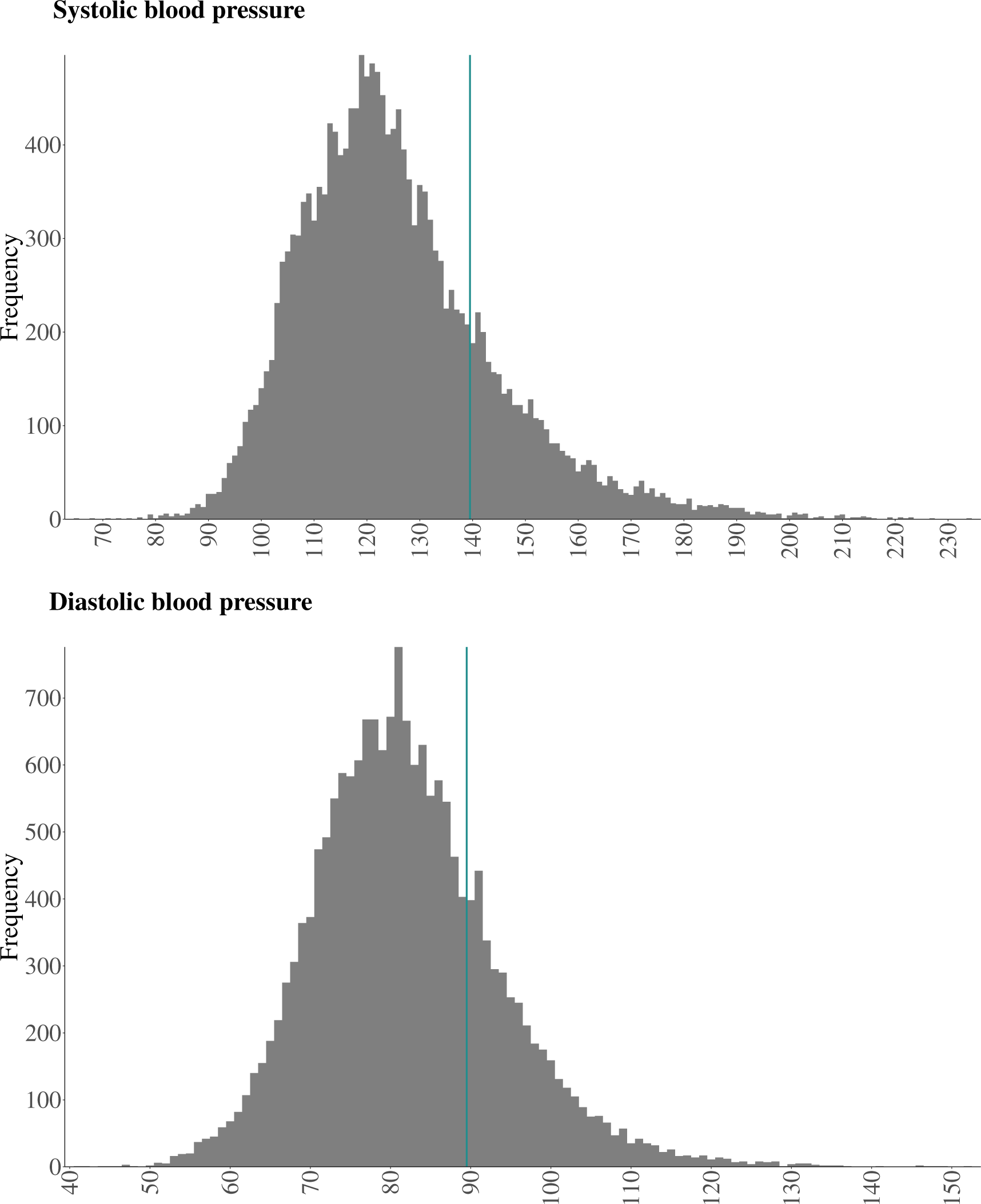
Frequency of each systolic and diastolic BP measurement result among adults aged 30 years or older Note: The vertical line in the top panel marks the threshold at a systolic BP of 140 mmHg and in the bottom panel at a diastolic BP of 90 mmHg.

